# Assessing the heterogeneity in the transmission of infectious diseases from time series of epidemiological data

**DOI:** 10.1101/2022.02.21.22271241

**Authors:** Günter Schneckenreither, Lukas Herrmann, Rafael Reisenhofer, Nikolas Popper, Philipp Grohs

## Abstract

Structural features and the heterogeneity of disease transmissions play an essential role in the dynamics of epidemic spread. But these aspects can not completely be assessed from aggregate data or macroscopic indicators such as the effective reproduction number. We propose an index of effective aggregate dispersion (EffDI) that indicates the significance of infection clusters and superspreading events in the progression of outbreaks by carefully measuring the level of stochasticity in time series of reported case numbers. This allows to detect the transition from predominantly clustered spreading to a diffusive regime with diminishing significance of singular clusters, which can be decisive for the future progression of outbreaks and relevant in the planning of containment measures. We evaluate EffDI for SARS-CoV-2 case data in different countries and compare the results with a quantifier for the socio-demographic heterogeneity in disease transmissions in a case study to substantiate that EffDI qualifies as a measure for the heterogeneity in transmission dynamics.

## 1 Introduction

Tipping points that define significant transitions in the infection dynamics or ramifications of an epidemic have been a main focus of attention for researchers, decision makers, media outlets and the general public during the ongoing COVID-19 pandemic. In particular decision makers often cite thresholds for indicators such as the number of intensive care patients, the seven-day case rate, the hospitalization rate or the effective reproduction number either as goals or justifications for containment measures. Most time-varying indicators that are commonly used to monitor an ongoing outbreak only measure the immediate effects of the outbreak on the population or, in the case of the effective reproduction number, aggregate properties of the current infection dynamics from a macroscopic point of view. However, outbreaks that are strongly governed by large infection clusters and superspreading events (SSEs) can only be fully understood by taking into account events occurring at a meso- or even microscopic scale of the epidemic. Such outbreaks are predominantly investigated with individual- or network-based models [2, 4, 8, 44, 46–48, 50, 51] and usually exhibit a large dispersion in the number of individual secondary infections, which has been observed to crucially affect the respective infection dynamics [2, 4, 13, 15, 34, 47, 48].

Specifically during low-prevalence periods of an epidemic, such as in the onset of an outbreak, a strong variation in the number of individual secondary cases implies a higher degree of stochasticity in the observed daily case numbers and increases the likelihood of stagnation as compared to a situation in which the offspring distribution exhibits only a small variance [4, 34]. Once an outbreak with large dispersion in the individual secondary case numbers has taken off and grown beyond the emergence of isolated SSEs, it usually exhibits stable exponential growth with growth rates that are comparable to an outbreak with the same basic reproduction number but no variation in the number of secondary infections per infected individual [4, 34]. This suggests a phase transition occurring in the early stages of an outbreak with large dispersion in the number of individual secondary infections after which containment becomes increasingly difficult due to the ill nature of exponential growth. Correctly observing this tipping point in the course of an ongoing outbreak would thus be of great value for any evaluation of its time-varying infection dynamics.

In this work, we propose a novel indicator that makes this phase transition transparent by quantifying the *effective aggregate dispersion* of epidemic outbreaks based on time series of daily reported case numbers and a statistical model for reproduction. Technically, our indicator can be seen as a time-varying measure for the stochasticity in time series of aggregate daily case numbers. It is important to note that simply computing standard measures of variation, such as the empirical variance, does not yield a meaningful metric in this setting. This is mainly because much of the variation is not due to the dispersion of secondary infections but can be explained by other factors like changes in the effective reproduction number or weekly periodic patterns that are caused by seasonalities in the behavior of the population and the underlying testing and reporting regime. Our statistical model and inference framework take into account artifacts and patterns that cannot be attributed to the dispersion in the number of secondary infections and provides an *effective aggregate dispersion index* (EffDI) that reflects the heterogeneity in individual transmission dynamics. We anticipate the EffDI to act as an ‘early warning system’ that allows decision makers to react to subtle but significant changes in the infection dynamics of an outbreak during periods of low-prevalence.

Investigating the infection dynamics on the individual or mesoscopic level furthermore requires a model for reproduction that considers the temporal aspects of disease transmission and case registration that manifest in aggregate reported case numbers. We revise in subsection 2.1 and subsection 2.2 the structure of existing models for inferring effective reproduction factors [1, 11, 20, 22, 31, 39, 40, 49, 51] and basic dispersion parameters [2, 4, 11, 13, 15, 34, 47, 48] and generalize reproduction as the interplay between *infectious load*, which corresponds to the current contagious population that can be derived from reported case numbers, and *infectious activity*, which is the corresponding amount of new infections. We then analyze in subsection 2.3 the characteristics of the stochasticity in time series of reported case numbers and artifacts that result from the administration and procedures of case registration. Based on our investigations, we develop in subsection 2.4 a carefully designed infection model for the quantification of time-varying effective aggregate dispersion in daily incidence time series. This model relates the infectious load observed at a given day with the respective infectious activity and has certain desirable properties that enable the quantification of time-varying effective aggregate dispersion. Most importantly, it is capable of capturing daily seasonal patterns in the reproduction dynamics and considers an additional time-varying aggregate dispersion parameter. Based on this model, the EffDI will be determined by the minimal amount of time-varying aggregate dispersion required such that the observed pairs of infectious load and infectious activity during a predefined time window are plausible under the fitted model parameters.

With our approach, we exploit the connection between the observed stochasticity in reported case numbers and the degree to which SSEs and infection clusters are statistically relevant for the current infection dynamics. To support the developed indicator, we investigate in subsection 2.5 the occurrence of SSEs from a socio-demographic perspective based on reported case data that includes information about a small number of social attributes of the incidence population. We measure the statistical distance between the socio-demographic composition of infectious load and infectious activity, and assume that this supporting indicator reflects the emergence of infection clusters that are significant relative to the overall infectious activity. From the qualitative correspondence of both indicators, we conclude that EffDI can indeed be used to assess the significance of SSEs and infection clusters based on a time series of aggregate reported case numbers.

## 2 Results

### 2.1 Inference of reproduction dynamics

The common approach for investigating the reproduction dynamics of epidemic outbreaks aligns with a mathematical model that can be formalized as

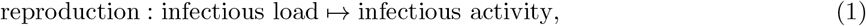

where *infectious load* refers to the currently contagious population or the induced potential for generating new infections among the susceptible population, and *infectious activity* refers to the amount of infections arising from transmissions by the currently infected population.

For inferring information about reproduction, load and activity are regarded as ‘exogenous’ variables that are derived from reported case numbers. This transformation must take into account the characteristics of the disease and of case reporting. An important simplification is to assume that load and activity are equally affected by under-reporting and by gradual changes of the detection rate. This becomes evident for a simple multiplicative reproduction factor in particular. Nevertheless, registration of cases is also subject to specific delays which result from the deferred exposure of symptoms, personal testing and monitoring schemes, and from the implemented administrative processes (compare ‘nowcasting’) [1, 24, 28, 38, 42]. Furthermore, to distinguish between load and activity, the time between subsequent infections in a transmission pair must be considered [10, 18, 20–22, 25, 29]. This time period is specific to the disease and the contact behavior of the population and can only be measured in individually tracked infection pairs [3, 6, 14, 23, 32, 33, 36, 37]. Figure 1A presents a schematic diagram of the time intervals occurring in transmission pairs. Figure 1B visualizes the general approach for inferring reproduction via load and activity from reported case numbers.

In the following, we investigate the statistical distributions of typical SARS-CoV-2 disease intervals and formalize the calculation of load and activity (subsection 2.2). We then analyze the stochasticity and artifacts in time series of reported SARS-CoV-2 case numbers (subsection 2.3) which is the foundation for constructing a statistical framework for the quantification of heterogeneity in reproduction in the form of an aggregate dispersion parameter (subsection 2.4).

**Figure 1:**
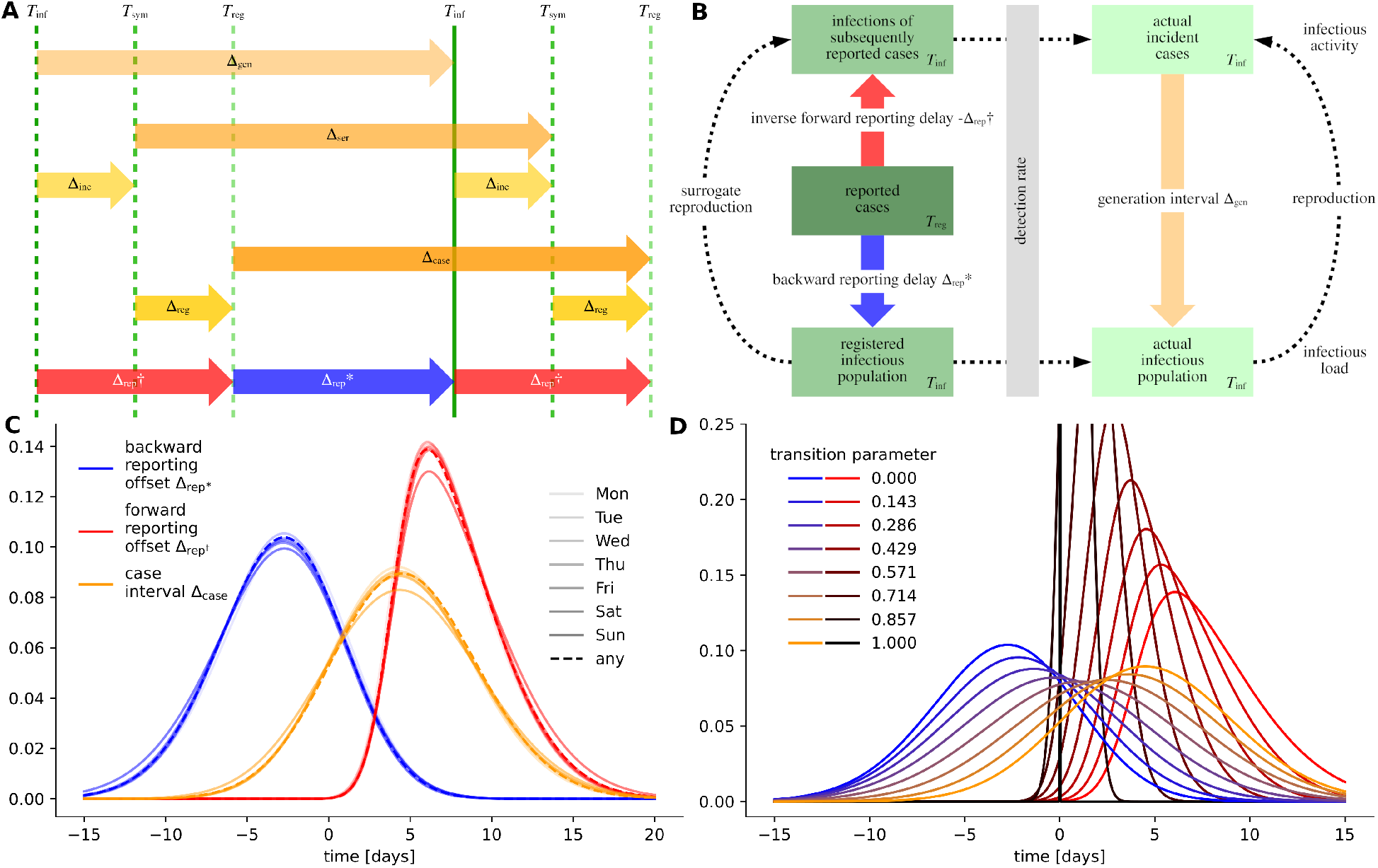
(A) Disease and transmission intervals. Schematic diagram showing the events *infection, symptom onset* and *registration* (denoted by *T*) in the timeline of a hypothetical infection pair. The time period between two consecutive disease transmissions is called the *generation interval*, here written as Δ_gen_. The time period between the symptom onsets in an infection pair is called the *serial interval* Δ_ser_. We further denote the time period between registration of two consecutive cases as the *case interval* Δ_case_. Further intervals shown are the *incubation delay* Δ_inc_, the *registration delay* Δ_reg_ and the *forward and backward reporting offsets* 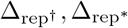. (B) Visual outline for the quantification of reproductive dynamics based on time series of reported cases. Infectious load and activity can be derived from reported cases using the statistical distributions of the reporting offsets. The obtained characterization of epidemic progression is only a surrogate for actual reproduction dynamics. (C) Probability densities of the backward and forward reporting offset distributions and the case interval distribution inferred from data. All interval distributions were calculated for different parameter settings; here, the weekday of case registration is encoded in the lightness of the color. (D) Transformation between statistical models for the reporting offsets. The statistical models inferred from data can be continuously transformed, for instance, into a delta distribution and the case interval respectively.

### 2.2 Obtaining infectious load and activity

We denote the time interval between infection and (potential) registration in a surveillance system as the *forward reporting offset* 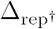, and the time interval between the registration of a case and a (potential) secondary infection as the *backward reporting offset* 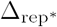. The *case interval* 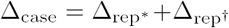 is the time between the registration of consecutive cases. Independently of the actual statistical distributions of these time intervals, load and activity can be interpreted as simulated contagion events or as the corresponding time series,

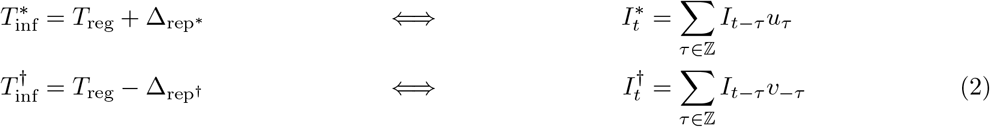

where *I*_*t*_ is the original time series of reported cases, which is composed of registration events *T*_reg_, and *u*_*τ*_ and *v*_*τ*_ are the probability masses of the backward and forward reporting offset distributions. Hence, the time series of infectious load 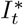 can be understood as the collection of the events 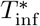 sof persons (potentially) transmitting the disease and the time series of infectious activity 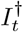 represents the events of persons getting infected 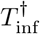.

To obtain a tangible statistical model for the temporal aspects of contagion on the individual level, we formulate an algebraic equation system that relates the time intervals occurring in transmission pairs (compare Figure 1A). Integrating data about the registration of cases, we then ‘solve’ this equation system using a Markov chain Monte Carlo (MCMC) approach (see *Methods*, subsection 4.1). To account for the most significant temporal changes and seasonalities in reporting, we separately perform our calculations for different stratifications of Austrian SARS-CoV-2 data [27] differentiating by the day of the week and for distinct time periods of the epidemic. Corresponding with the availability of testing capabilities, we observe that in the early stage of the pandemic and on weekends, the delay between infection and registration is longer (Figure 1C). In general, the inferred interval distributions exhibit slightly increased variance compared to the intervals found in literature. Detailed results and comparison is provided in Supplement 8. On average, without distinguishing different phases or weekdays, we find probability distributions with the mean values and standard deviations 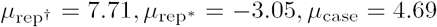 and 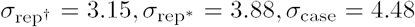. The obtained statistical models are particularly valid for the Austrian setting with a relative large number of routine tests, but could indicate a general configuration that is also applicable to other countries. Similar approaches based on MCMC methods or Bayesian inference were applied for synthesising missing statistical information about disease intervals before [1, 9, 19, 21, 24, 36, 49].

In practice, frameworks that deal with the quantification of reproduction factors often use – in contrast to (2) – a simplified model of disease and transmission intervals to calculate what we call load and activity. A reason for this is that particularly during the early stage of a pandemic detailed knowledge about the time intervals, and especially the reporting dynamics, is not available [11, 40, 49]. Often – partially justified by the characteristics of the observed disease – estimates of the serial interval provided by secondary literature and studies are used to approximate the generation or case interval [4, 11, 40, 51]. Methods for extracting the relevant inter-patient time distribution from a number of observed infection pairs were also directly included in the algorithms for calculating reproduction factors [49]. Furthermore, in the inference of reproduction factors, reported cases are widely used as a surrogate for actual contagion events [11, 16, 20, 22, 30, 40, 49, 51] (infectious activity). The corresponding ‘simplified’ configuration of the interval model and the associated forms of infectious load and activity corresponds to the formula

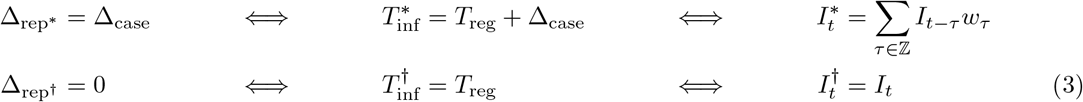

where *w*_*τ*_ are the probability masses of the distribution of the case interval Δ_case_, which is usually replaced with either Δ_ser_ or Δ_gen_. Under the condition that the case interval – or the used surrogate – is strictly positive, it can further be argued, that in contrast to ‘complex’ data-driven models (as discussed above) using (2), this form is better suited for the real-time assessment and forecasting of epidemics as no future values in the time series of the reported cases is required [11, 20, 22, 49].

### 2.3 Stochasticity in reported case numbers

Distinct properties of reported (SARS-CoV-2) case data are weekly seasonal patterns [13, 24, 31, 38, 39] and artifacts that result from reporting procedures. Furthermore, the general level of stochasticity is high in low-incidence phases but diminishes when case numbers become large, rendering the data more regular with pronounced seasonal patterns in these periods. Figure 2 illustrates this phase separation for three different countries by comparing normalized segments of reported case numbers from low-prevalence periods with segments from high-prevalence periods.

**Figure 2:**
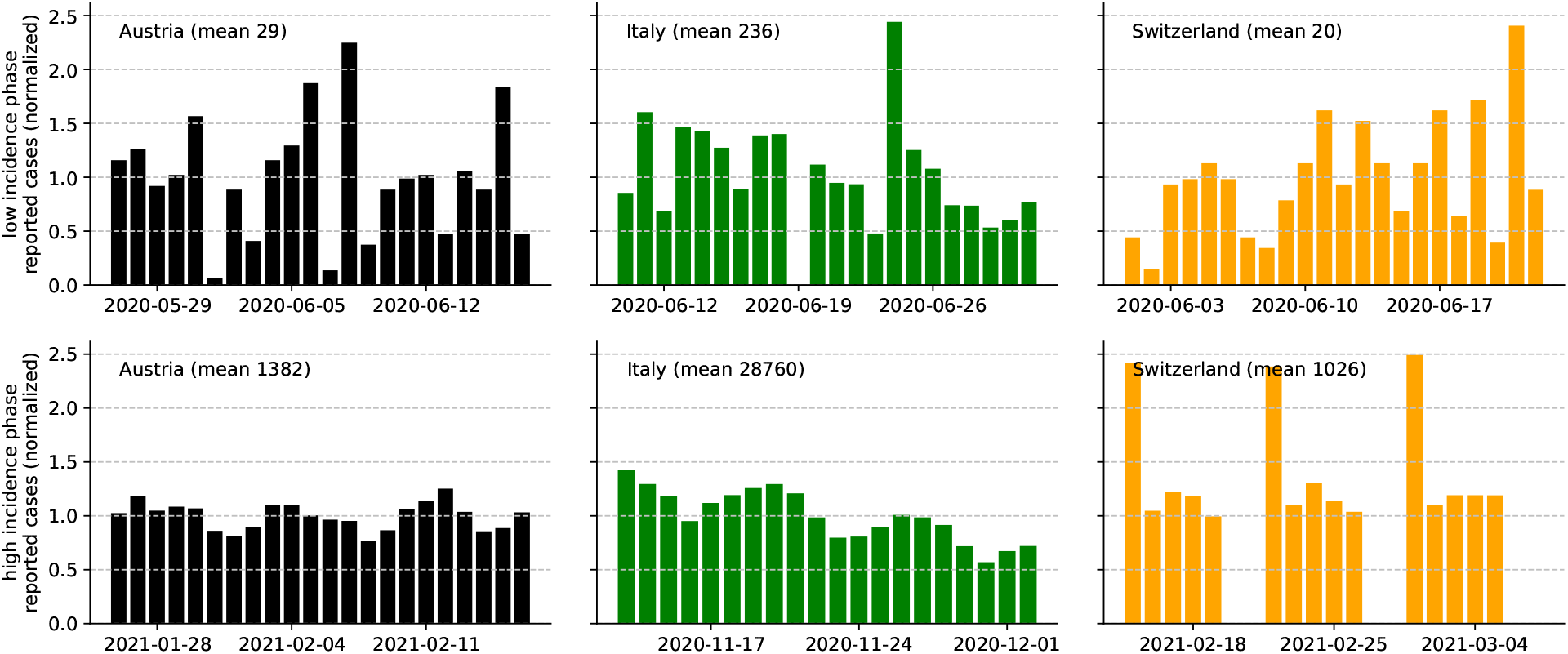
Normalized segments of daily reported case numbers [12] during low-prevalence (top row) and high prevalence (bottom row) periods in different countries (columns). Normalization was achieved by dividing the daily case numbers by their mean value during the respective three-week period. During the respective high prevalence periods, we observe a high degree of regularity and weekly seasonal patterns, whereas in low-prevalence phases, the segments seem to be more erratic.

Statistical models for inferring effective reproduction factors usually directly operate on crude reported case numbers according to the model in (3). One way to account for the at times large variance in the data is to increase the dispersion of the statistical model for the individual number of secondary infections (offspring distribution). In other words, when the data is over-dispersed with respect to the employed statistical model, a model with greater variance in secondary infections must be used. Alongside improved correspondence with the data [4, 30, 35], such a modification can also provide – eventually in the form of a *basic dispersion parameter* – information about the inherent level of dispersion in the secondary infections during an outbreak [2, 15, 34, 47]. This, in turn, is a leverage point for the assessment of spreading characteristics such as the occurrence of SSEs [2, 4, 34, 48]. However, the stochasticity and regularity in time series of reported case numbers is not only affected by the transmission dynamics of the epidemic. It is especially the employed testing and reporting regime that causes the observed weekly seasonal patterns (Figure 2), making it necessary to separate artifacts of reporting from the stochasticity that is presumably caused by the transmission dynamics. Additionally, and also supported by the observations in Figure 2, it is a natural extension of the state-of-the-art methodology to consider a temporally varying degree of dispersion. Ultimately, this leads to what we call a time-varying *aggregate dispersion parameter*, which allows to differentiate periods of an epidemic that are driven by SSEs and episodes with diffusive spread. We implement both aspects in our extension of the existing statistical inference framework in the next section (subsection 2.4).

The straight-forward approach for pruning time series of reported case numbers of artifacts and noise is by smoothing. This is essentially implemented by the ‘regularizing’ model defined in (2), which utilizes the previously inferred offset distributions as convolution kernels [1, 20, 31]. Notably, with this approach, besides regular patterns resulting from case reporting, also the roughness resulting from transmission dynamics is filtered out to a certain extent, such that evidence about the stochasticity in transmission dynamics is lost. We anticipate, that the ‘raw’ form of load and activity (3) is more suitable for harnessing the stochasticity in reported case data [12] than a ‘regularizing’ model (2). In Supplement 12 we further address the question of how much of the stochasticity in reported case data can actually be filtered out in the calculation of load and activity before the assessment of stochasticity fails. We investigate the quantifiers developed in this paper under a continuous transition between the ‘pre-regularizing’ model in (2) and the ‘raw’ scenario defined in (3) (cf. Figure 1D). Our results confirm what has been observed in existing research [7, 11, 20, 22, 49]. Regularization, on the one hand, leads to implicit delays in the time series of load and activity, which is problematic in the real-time assessment of epidemic progression. Secondly, over-smooth input time series can hinder the assessment of stochasticity. On the other hand, when reporting artifacts or excess roughness in the underlying data is not filtered, the confidence intervals in statistical inference approaches are generally larger and numerical quantifiers can behave erratically. Furthermore, we conclude that if reporting artifacts are sufficiently filtered, inclusion of detailed data-driven statistical distributions of disease and transmission specific intervals can be evaded [20, 30, 31] and it is reasonable to map only the basic characteristics of these intervals.

### 2.4 A model for quantifying effective aggregate dispersion

A common statistical approach for the inference of reproduction dynamics is based on a Poisson model for the individual number of secondary infections (offspring distribution) that is parameterized with a reproduction factor as the rate or expected value. Under the assumption that the individual reproduction factor (i.e. the expected number of secondary cases) is the same for all contagious individuals at a given time *t* and by using the additivity of the Poisson distribution, the total number of new cases 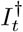 (infectious activity) can be modelled as

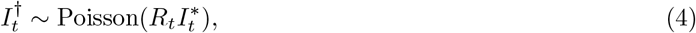

where 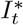 is the number of contagious individuals (infectious load), and *R*_*t*_ denotes the common reproduction factor [11, 40, 49].

Not limited to the current SARS-CoV-2 pandemic, a large variety of techniques and software packages have been developed to infer time series of aggregate reproduction factors *R*_*t*_ based on (4). A particular extension of this approach is to regard the individual reproduction factor as a random variable instead of a deterministic uniform value. Typical models for a stochastic individual reproduction factor are for instance the exponential distribution, or the gamma distribution. Due to the stochasticity encountered in aggregate reported case numbers, using an exponential model can lead to over-dispersion when fitting the resulting geometric model to the data [31, 34]. Considering identically and independently gamma distributed individual reproduction factors with shape parameter *k* and scale parameter *R*_*t*_*/k* leads to a negative binomial model (gamma-Poisson mixture) for the total number of new cases, which can be written as

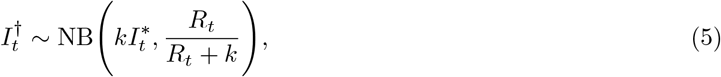

where the first parameter is understood as the number of allowed failures and the second parameter is the success probability. This approach allows the model to better comply with the observed stochasticity in reported case numbers [2, 4, 24, 30, 31, 34, 35] by considering an additional free parameter *k*, which can be viewed as a dispersion parameter that reflects the variance in individual secondary cases. Large dispersion, which is associated with a scenario that shows great variability in the individual numbers of secondary cases, can be modelled in (5) by choosing *k* small. Vice versa, small dispersion, which describes a situation where all infected individuals cause a similar number of secondary cases and where spread of the disease happens in a more ‘homogeneous fashion’, can be modelled by choosing *k* large. For *k* → ∞, the limiting distribution falls back to the Poisson model (4). The properties of the negative binomial distribution proved to provide good overall correspondence with the data in many case studies. Analogous to the basic reproduction number, *basic dispersion parameters* of SARS-CoV-2 outbreaks and other epidemics have been estimated from historical data and used for investigating the heterogeneity of infection dynamics [2, 15, 34, 47, 48].

The observed variability of relative stochasticity in recorded time series data (Figure 2) and formal considerations regarding the model defined in (5) indicate that the effect of dispersion on the infection dynamics is negligible in high-incidence periods. In order to capture the variable impact of dispersion on the dynamics of an outbreak with a quantitative measure, we consider a carefully designed infection model which introduces a time-varying *aggregate* dispersion parameter *κ*_*t*_,

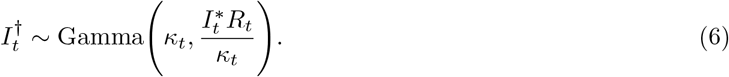

The key statistical aspects of the models (5) and (6) are recapitulated in Supplement 11. Essentially, both models have the same expected value 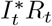, but the variance of the former scales linearly with 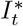, whereas the variance of the latter scales quadratically with 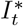. Hence, given that the true infection dynamics of an outbreak yield an infectious activity 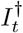 for which the variance scales subquadratically with the magnitude of the infectious load 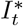 – as observed in the data (c.f. Figure 2) – we can choose the parameter *κ*_*t*_ in the model (6) increasingly large during high-incidence regimes without impeding the ability of the model to describe the variance present in the data.

In order to prevent strong oscillations of the inferred reproduction number *R*_*t*_, inference approaches for existing models often assume constant reproduction during a certain time window [11]. To further remove the contribution of reporting artifacts and seasonal patterns to the observed variance, we describe the effective reproduction number *R*_*t*_ in (6) with a linear model that takes into account trends and weekly periodic patterns. Ultimately, for a given day *t*, we apply Monte Carlo simulation to estimate the smallest plausible amount of aggregate dispersion that is required for explaining the observed time series of daily aggregate case numbers.

Finally, we define the *effective aggregate dispersion index* (EffDI) in terms of the largest value for the time-varying aggregate dispersion parameter *κ*_*t*_ (i.e., the least amount of dispersion) for which the model (6) is plausible given the observed data around day *t*. Let *p*_*t*_(*κ*_*t*_) denote the plausibility of the parameter *κ*_*t*_ around day *t* (cf. (20)) and *κ*_*t*_(*p*) the largest value for *κ*_*t*_ such that *p*_*t*_(*κ*_*t*_) is greater or equal than a threshold *p* ∈ [0, 1]. Then, the EffDI on day *t* is defined as the square root of the reciprocal of *κ*_*t*_(*p*), that is,

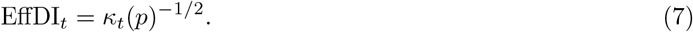

In this work, we usually set *p* = 0.9 and *p*_*t*_(*κ*_*t*_) is estimated for a fixed *κ*_*t*_ via Monte Carlo simulation. A detailed description of the complete framework operating on (6) and its derivation can be found in *Methods* (subsection 4.2).

Figure 3B depicts over the timeline the plausibility of a range of dispersion parameters *κ*_*t*_ when fitting the corresponding model (6) on aggregate reported SARS-CoV-2 case numbers in Austria [12]. The smoothed approximation of the level set for *p* = 0.9 marks the abrupt transition from fully plausible dispersion parameters to model configurations for which the data is over-dispersed. This confirms that the aggregate dispersion parameter *κ*_*t*_ can in general be chosen larger during high prevalence periods, and that low-prevalence periods of an outbreak are usually associated with a measurably higher degree of stochasticity relative to the current infectious load. For instance, we can observe a significant phase transition in the stochasticity of the time series of aggregate reported case numbers during the end of May and the first weeks of June in 2020.

**Figure 3:**
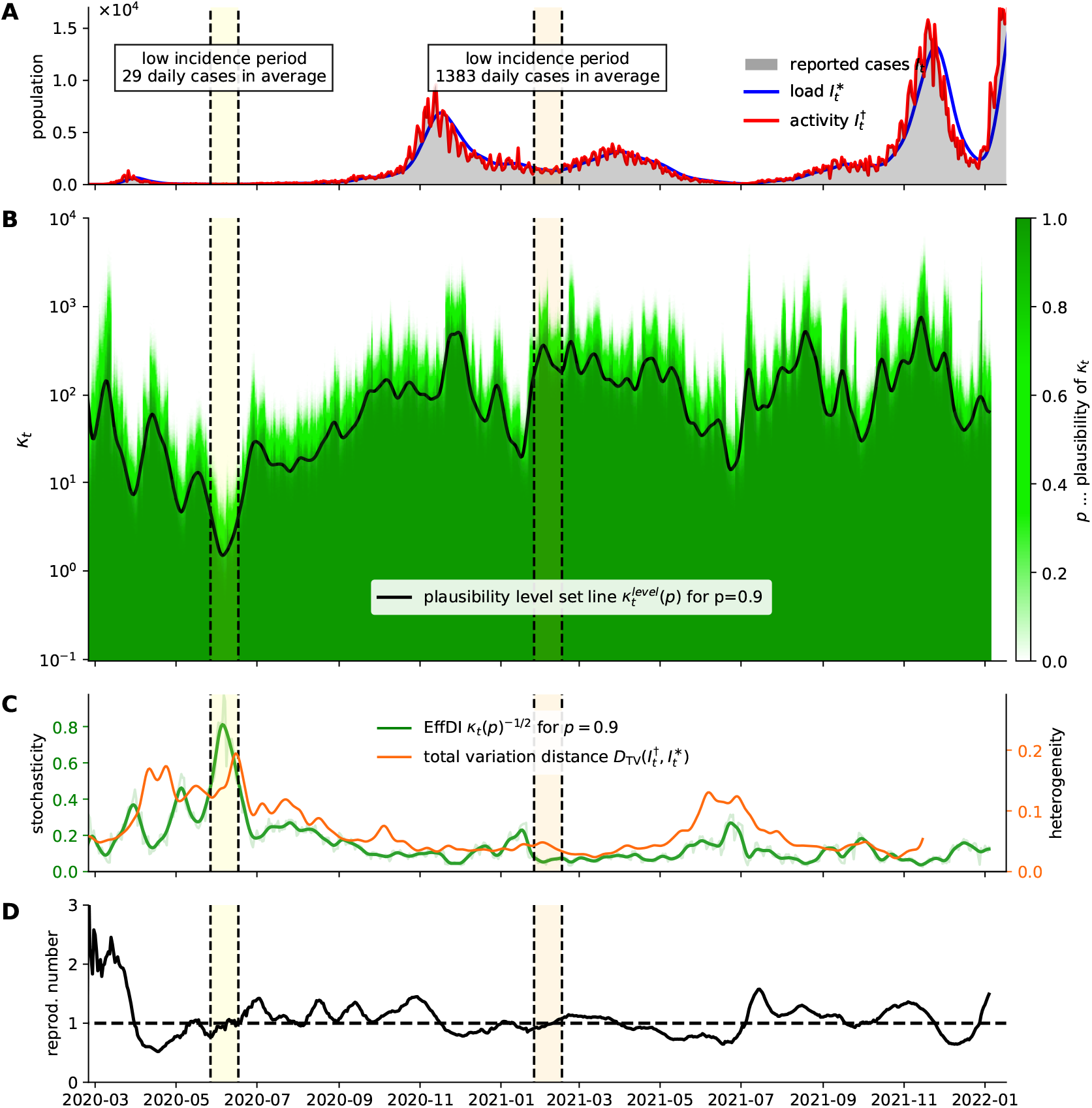
Quantification of effective aggregate dispersion in the reproduction dynamics of the SARS-CoV-2 epidemic in Austria [12]. (A) Number of reported cases and infectious load and activity according to (3). The low- and a high-incidence phases from Figure 2 are indicated with vertical lines. (B) Plausibility diagram for the dispersion parameter *κ*_*t*_. The transition between the plausible in implausible regime (e.g. *p* = 0.9) is abrupt. (C) EffDI results by the transformed level set line (green) (*κ*_*t*_(*p* = 0.9))^−1*/*2^. It is a quantifier for dispersion and corresponds to the coefficient of variation in the underlying statistical model (6). The progression of dispersion aligns with the progression of a socio-demographic heterogeneity measure (orange). Socio-demographic heterogeneity is measured via the total variation distance between infectious load and infectious activity (subsection 2.5). (D) Resulting case reproduction number.

In Figure 3C, we compare the EffDI with the progression of a measure for the socio-demographic heterogeneity in disease transmissions for the Austrian setting. The heterogeneity measure relies on data that includes additional information about cases and is developed in the next section (subsection 2.5). We motivate this measure as an independent indicator for the occurrence and significance of SSEs and infection clusters. As a consequence, qualitative correspondence of both measures substantiates that the progression of the time-varying aggregate dispersion parameter adequately reflects structural changes in the underlying infection dynamics of an outbreak.

Figure 4 shows the development of EffDI based on daily aggregate case numbers between February 25, 2020 and January 31, 2022 for six different countries. In the case of South Korea, the transition line closely follows the expected behavior of high stochasticity during low-prevalence periods and low stoachsticity during high-prevalence periods with a prominent phase transition during a particular period of low-prevalence around the beginning of May 2020. By far the longest period of high stochasticity can be found in the aggregate case numbers reported by Singapore, which yield a very high stochasticity measure for more than a year. In all of the five analyzed European countries (Austria, Switzerland, United Kingdom, Italy, and Germany), significant periods of high stochasticty can be observed between May and July 2020, indicating that during this time, the infection dynamics of the COVID-19 outbreak were mainly governed by comparatively few isolated events throughout many countries in Europe. However, in particular in the cases of Germany and Italy, our analysis also yields periods of high stochasticty during times of high prevalence. In these cases, it is not clear whether the observed stochasticity in the aggregate case numbers should be attributed to the actual infection dynamics or changes and inconsistencies in the employed testing and reporting regime. A particular country where basically all periods of high stochasticity in times of high prevalence can in fact be attributed to changes and irregularities in the reporting is Switzerland, where aggregate case numbers were only reported for weekdays after September 19, 2020, and where four additional dates in the spring of 2021 were missing from the considered time series.

**Figure 4:**
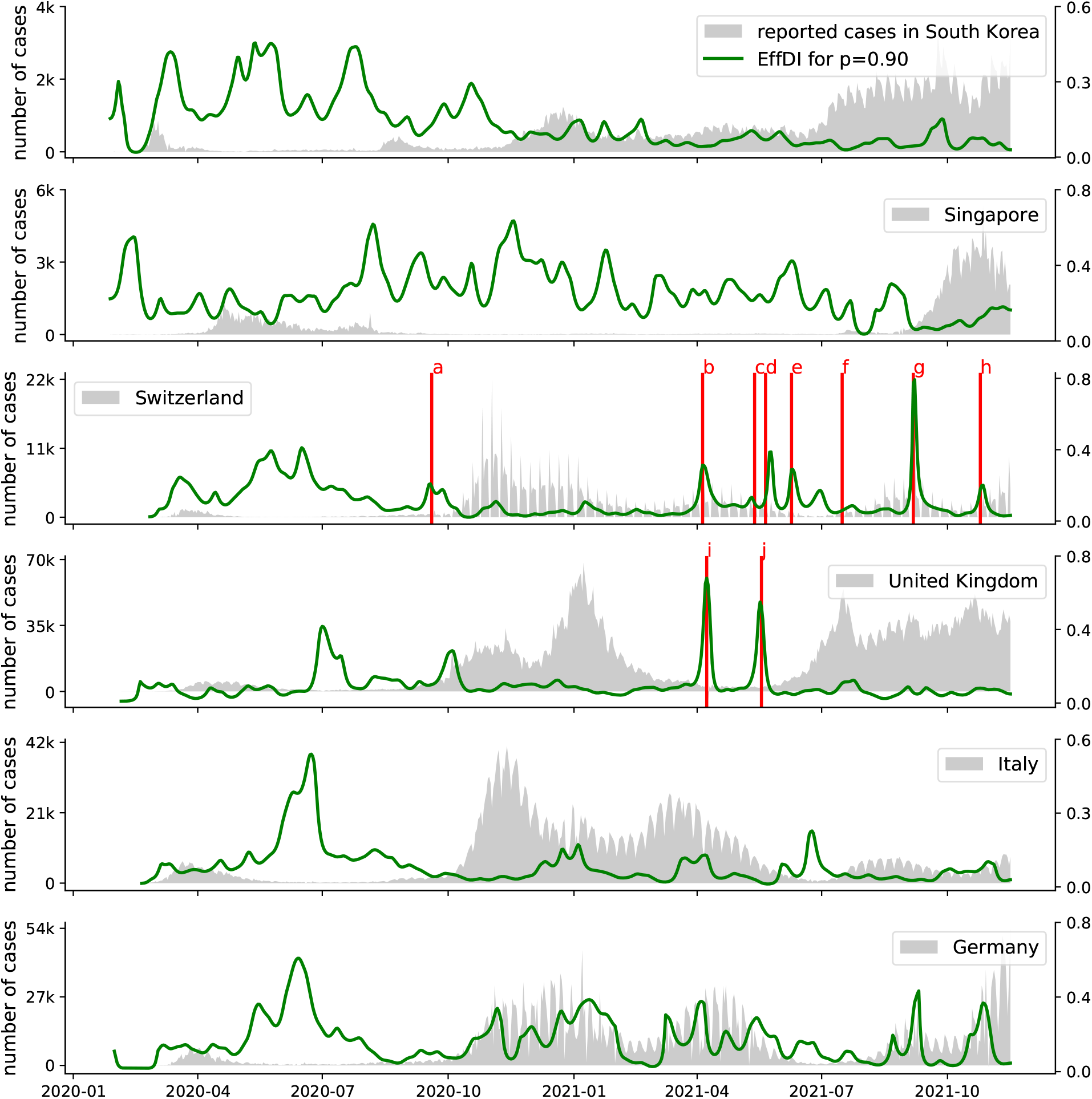
Evaluation of EffDI based on time series of daily aggregate case numbers for six different countries [12]. In the case of Switzerland and the UK, certain dates are highlighted to illustrate the effect of irregularities or changes in the reporting regime. a: After September 19, 2020, Switzerland only reported aggregate case numbers on weekdays (cf. Figure 2). b, c, d, e, f, h: Missing aggregate case numbers in Switzerland on several days in 2021. g: Missing entries on a Friday and the following Monday in Switzerland during September 2021. i, j: Reporting of negative aggregate case numbers on two days in April and May 2021 in the UK.

### 2.5 Social heterogeneity in transmission dynamics

To support our findings about the temporal variability of aggregate dispersion, we investigate the progression of socio-demographic heterogeneity of the infected population and in reproduction using statistical distance measures. In particular, we calculate the *total variation distance* (TV)

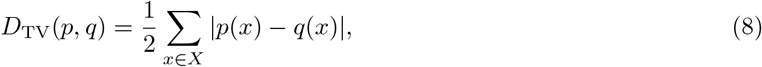

between normalized histograms *p* and *q* of two populations with respect to a discrete and categorical feature space *X*. Here *X* either refers to age-compartments, gender, geographic region or the product space.

Let 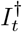 be the time series of vector-valued infectious activity consisting of normalized histograms of the newly infected population. Let *Q* be the socio-demographic configuration of the total population. We denote the difference 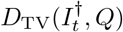 as the *heterogeneity of infectious activity*. If this quantifier is small, we expect that infectious activity is distributed homogeneously across the population. If the distance measure is large, infectious activity is concentrated in distinct social compartments. We use the data-driven interval models found in subsection 2.2 to extract vector-valued infectious activity according to (2) from an Austrian case data-set [26] and quantify the distance to the total population [5]. In Figure 5B-D the resulting heterogeneity of infectious activity with respect to gender, age and administrative affiliation is visualized. In Figure 5E the corresponding evaluated distance measures are plotted over time. With the visual as well as the quantitative approach it is possible to distinguish phases with heterogeneous infectious activity and phases with homogeneous activity.

**Figure 5:**
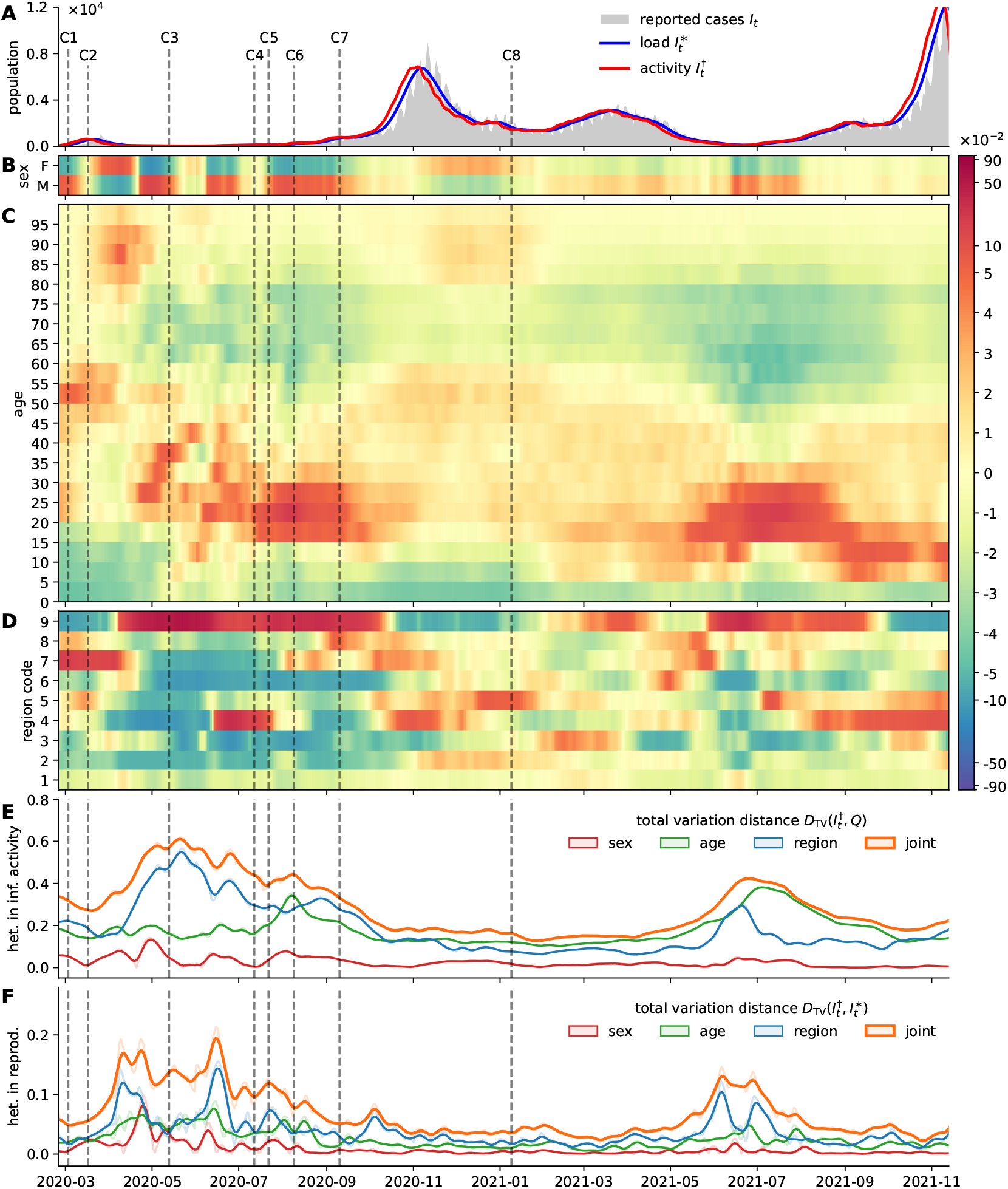
Heterogeneity of infectious activity and in transmissions inferred from reported case data in Austria [5, 26]. For calculating load and activity according to (2), the disease interval models presented in subsection 2.2 were used. Media coverage about certain SSEs is indicated by vertical dashed lines and the identifiers C1-C8 (see main text). (A) Crude number of reported cases; estimate for the time series of infections of subsequently reported cases (infectious activity); and estimated time series of the currently infectious population (infectious load). (B-D) For the social dimensions sex, age, and geographic region, the difference of the distribution of the estimate newly infected population (infectious activity) to the distributionof the total population 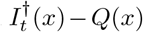 is visualized. High values (red) indicate over-representation of infectious activity and low values (blue) indicate under-representation. (E) Quantification of the heterogeneity of infectious activity using the total variation distance measure. To increase lucidity, a smoothed version of the resulting time series is shown. (F) Quantification of the heterogeneity in reproduction; if the distance measure is small, spread is confined to specific social strata; if the distance is large infections shift to previously unaffected social compartments.

We further compare the socio-demographic configuration of infectious activity 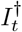 with the configuration of infectious load 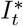 and denote the progression of the statistical distance 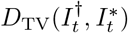 as the socio-demographic *heterogeneity in reproduction*. When the distance measure is small, then the socio-demographic configuration of the infected population remains the same, when the distance measure is large, we expect that the socio-demographic configuration of the infected population is about to change. Hence, large heterogeneity can indicate that predominant infectious activity shifts to previously unaffected social strata. We further assume that abrupt changes in the socio-demographic composition of the infected population point to the emergence of infection clusters that are composed of individuals with similar social attributes (e.g. geographic location). Hence, we conclude that a measure for the heterogeneity in reproduction can indicate the emergence and dissipation of significant infection clusters. In Figure 5F this measure is evaluated for Austrian case data [26] using the interval models from subsection 2.2.

In Figure 5 a selection of significant SSEs and infection clusters is indicated in the timeline (compare [2, 4]). Details about case numbers and the corresponding media coverage are found in the supplement (Supplement 10). We observe an initial cluster (C1) in Ischgl, Tyrol (region code 7) affecting mostly younger and middle-aged persons. During the first epidemic wave, news media report about infection clusters in retirement homes in different provinces (C2). During the summer of 2020 we further observe mostly smaller infection clusters with occasional larger SSEs (C3) in logistic centers in and around Vienna (region code 9) as well as other SSEs (C4) in the state of Lower Austria (region code 3). A large regional cluster (C5) was detected in Upper Austria (region code 4). In the onset of the second epidemic wave SSEs are reported to have occurred during private meetings (C6) as well as in public events (C7). After a high-incidence phase, infection clusters in retirement homes across the country were observed (C8). Media reports about specific (noticeable) SSEs occurred in particular during the low-incidence phases of the epidemic. Analogously, the measure for heterogeneity in infectious activity is large only during the same periods indicating that individual SSEs can be distinguished only during such phases of an epidemic. In high-incidence phases, singular SEEs can be recognized in measured heterogeneity only if they are very significant in size (relative to the current incidence numbers) involving hundreds of infections.

## 3 Discussion

The methods proposed in this paper aim to provide insight into the mesoscopic dynamics of epidemic outbreaks based on time series of reported case numbers. A statistical framework was designed to harness the stochasticity in such data for inferring a time-varying *effective aggregate dispersion index* (EffDI) that reflects the progression of heterogeneity in the configuration of individual secondary infections. Large dispersion is in general associated with clustering and the occurrence of superspreading events (SSEs), whereas low dispersion implies homogeneous reproduction and spread [2, 4, 34, 35, 47, 48]. Technically, the proposed indicator quantifies the time-varying minimal plausible amount of dispersion that is required for explaining the stochasticity in observed incidence numbers via a statistical model for reproduction. As a consequence, this novel approach serves to distinguish phases of an outbreak, in which infection clusters and SSEs are definite and play a significant role, from phases, in which infection clusters either appear indistinct or are nonexistent. Since the individual distribution of secondary cases is only indirectly reflected in our model, we use the notion of a dimensionless *aggregate* dispersion parameter to emphasize the absence of a direct ‘physical’ interpretation. Correspondingly, the proposed indicator does not quantify the (plausible) variance in individual secondary infections but rather measures the plausible heterogeneity of the spreading dynamics on a mesoscopic scale. For instance, we assume that EffDI also accounts for the occurrence of multi-generation clusters, instead of assessing the likelihood for individual superspreaders.

Evaluating the EffDI for a number of countries we observe that the transition from periods with moderate case numbers to periods with high incidence (‘epidemic waves’ or ‘peaks’) is often accompanied by the early decline of the EffDI. We speculate that this behavior indicates a pending shift in the quality of infectious spread towards the diffusive regime with exponentially growing case numbers. Hence, the EffDI could be a valuable tool for monitoring qualitative changes in the mesoscopic spreading behavior. We anticipate that our novel indicator will be relevant in the planning of containment measures (e.g., deciding between individual-level contact tracing policies and large-scale containment measures) [4, 13, 47, 48, 50].

Our approach is founded on a branch of existing statistical models for reproduction that have been developed for inferring the progression of effective reproduction factors [1, 11, 20, 22, 31, 39, 40, 49, 51] and a constant *basic dispersion parameter* [2, 4, 11, 13, 15, 34, 47, 48]. In alignment with the general concept of reproduction, the employed model portrays a relation between the currently contagious population (infectious load) and the new emerging infected population (infectious activity). For the extraction of both quantities from time series of reported case numbers, specific time intervals such as the incubation and generation period as well as delays in case registration must be considered. Furthermore, the implemented testing and reporting procedures often lead to weekly seasonal patterns and artifacts in the data. Experiments showed that modeling the seasonalities of case reporting in the statistical distributions of the disease intervals in a data-driven approach can successfully eliminate artifacts of reporting, but also obliterates the stochasticity, which presumably contains the effects of heterogeneous spreading dynamics. As a consequence we include a technique for the dynamic filtering of weekly periodic patterns within our statistical framework which allows to retain exactly the residual stochasticity of the data while at the same time correctly reproducing the relevant times intervals. Analogous to existing research [7, 11, 20, 31, 49], we confirm that in this setting there exists a trade-off between synthesizing load and activity based on ‘accurate’ data-driven models for disease and transmission intervals, and the careful abstraction of transmission dynamics.

We evaluate the progression of EffDI for reported SARS-CoV-2 case numbers in different countries and confirm that inferred effective aggregate dispersion is usually higher in low-incidence regimes. This can be explained by a generally higher relative stochasticity for small sample sizes [35] but also aligns with individual infection clusters only having a significant effect on the overall infection dynamic in such periods. Hence, EffDI can assess the presence and demarcation of infection clusters and SSEs in low-incidence periods, but in high-incidence periods a large number of simultaneous clusters can appear as homogeneous spreading and obliterate the significance of singular spreading events. Nevertheless, our indicator is not trivial in the sense that the time periods with elevated effective aggregate dispersion do not simply agree with periods of large relative variance in the data, constantly low case numbers or a large effective reproduction factor.

To further validate the proposed indicator, we use an Austrian SARS-CoV-2 data-set that includes aggregated social attributes of reported cases and compare the progression of socio-demographic heterogeneity in disease transmissions with the progression of the detected effective aggregate dispersion. Whereas our original method is data-economical, the validation approach relies on higher-resolution case data for quantifying the statistical distance between the socio-demographic configuration of the present and newly infected population (between infectious load and infectious activity). Typically, close-proximity interaction communities, which are the main driver for infection clusters and SSEs, are characterized by distinct social attributes like age or geographic affiliation [2, 4, 13, 46–48]. Hence, we assume that the demarcation and significance of individual infection clusters and SSEs can be deduced from relatively abrupt changes in the observed socio-demographic composition of the incidence population. The indicators show good qualitative correspondence and we can substantiate that EffDI is suitable for differentiating regimes with clustered spreading characteristics form regimes with diffusive spread.

The EffDI complements existing indicators (e.g., effective reproduction factor) which are used for assessing and anticipating the dynamics of epidemic outbreaks. Furthermore, the separation and indication of periods with different spreading characteristics could also improve individual-based [8, 41, 44, 47] and aggregate models for the simulation and forecasting of epidemics and could provide a reference for combining or switching between different modeling approaches. We also assume that providing indications about the time-varying progression of heterogeneity in epidemic outbreaks could contribute to the general perception of epidemic spreading as complex dynamics on multiple scales. Generally, the usability of quantifiers and numerical indicators is inevitably affected by the availability and accuracy of data. Especially in the epidemiology of infectious diseases, the data often provides an indirect view on the actual situation and makes it necessary to combine data of different origin and to map the dynamic aspects of spreading by the means of mathematical models. An immediate consequence is to include also the variability or stochasticity of reported case data itself in such models and in the calculation of numerical indicators. For instance, in the inference of effective reproduction factors, the relative stochasticity of the data is used for assessing the uncertainty of statistical estimates [11, 30, 49]. Analogously, we designed our approach to use the stochasticity in the data for inferring information about the infection dynamics on a mesoscopic scale, that can not directly be concluded from aggregate case numbers. However, for investigating the plain stochasticity or regularity in time series of reported case numbers there exist more suitable mathematical approaches like approximate entropy or spectral methods that do not integrate a statistical model for reproduction.

We provide the algorithms developed in this paper as a Python programming package (Code availability) that can operate on any time series of crude reported case numbers. We anticipate that the proposed concept of an indicator for effective aggregate dispersion can further be improved in terms of the reflection of artifacts and seasonal patterns in the data and by investigating the robustness against changes in the assumed characteristics of the disease. In our current approach, abrupt changes in the seasonality of the data, which are caused by systematic changes in case reporting operations, can lead to the artificial inflation of inferred effective aggregate dispersion. Furthermore, our framework does currently not regard truncation or other aspects that should be considered in the analysis of time series data. Besides technical improvements, we assume that investigating effective aggregate dispersion with synthetic data that was obtained from individual-based or network models [8, 41, 44, 46, 47, 50, 51] could provide additional insight and improvements.

## 4 Methods

### 4.1 Disease and transmission intervals

#### 4.1.1 Motivation

In the study of the epidemiology of transmissible diseases usually only a subset of the relevant time intervals is accessible via collected data. We relate an extended set of disease and transmission specific intervals in a stochastic model to infer information that cannot be retrieved directly from collected data in most cases.

Following subsection 2.2, we denote the offset between infection and (potential) case registration as the *forward reporting offset* 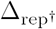 and the time between detection and a (potential and subsequently detected) secondary infection as the *backward reporting offset* 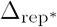. For an infection pair *AB* let 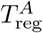 be the point in time when a patient *A* was registered, let 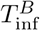 be the point in time when the secondary case (patient *B*) acquired the infection from patient *A* and let 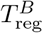 be the time of registration of patient *B*. Then

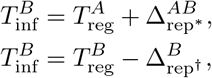

where 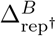 and 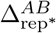 are the specific reporting offsets for the infection pair *AB*. In addition to the specific infection and reporting times *T*_inf_ and *T*_reg_, also regard the time of symptom onset *T*_sym_ and denote the time intervals

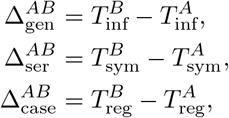

as the *generation, serial* and *case* interval. We further write

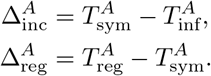

for the *incubation period* and the *registration delay* (for patient *B* respectively). A diagram relating all disease and transmission intervals is provided in Figure 1A.

#### 4.1.2 Data

For quantifying infectious load and activity (compare (2)), the time series of reported cases and the statistical distributions of the reporting offsets are required. In literature and studies, however, most often the generation [17, 21, 36] and serial intervals [3, 14, 25, 33, 37, 43] as well as the incubation period [6, 32, 33, 52] are investigated and the underlying data usually stems from a relatively small number of closely tracked infection pairs. Parameters of statistical distributions found in literature and visual comparison are provided in Supplement 1 and Supplement 2. In this stochastic inference approach we use ‘averaged’ versions of the found distributions.

Delays in registration and reporting, on the other hand, are less frequently studied since they are very specific to administrative processes that vary by country and have greater variability due to changing reporting procedures and laboratory schedules. We used a data-set provided to us by the Austrian Agency for Health and Food Safety [27] containing aggregated data about the time of symptom onset and of recording in a national surveillance system. We subdivided the data by registration date at 2020-06-01, 2020-10-01 and 2021-03-01 and extracted the registration delay distribution separately for each of the four resulting periods and for each day of the week. For each subset, the time between symptom onset and case registration was fitted with gamma distributions. Before fitting it was necessary to remove erroneous entries and certain outliers (delays longer than three weeks) and all entries with large negative delays (200 out of 300 000 entries until March 2021). We observe smaller delays in the later stages of the epidemic; presumably due to improved testing and registration procedures. Weekly patterns can be recognized with longer reporting delays for cases that first showed symptoms during the weekends. The obtained distributions are specific to the Austrian setting; we assume, however, that qualitatively similar shapes should be obtained for other countries. The inferred parameters are provided in Supplement 3. Visualizations of the data is provided in Supplement 4.

Population specific time intervals are less frequently studied than intervals in transmissions that are specific to the disease. As a consequence, in models for the inference of reproductive numbers, often the serial or generation interval is used as an approximation of the case interval [4, 11, 40, 51] (compare (3)). In fact, we find that 𝔼 [Δ_gen_] ≈𝔼 [Δ_ser_] ≈𝔼 [Δ_case_]. Nevertheless, the generation interval is always a positive time period, whereas the serial and case intervals can take negative duration. We distinguish statistical distributions for the serial interval found in literature that assume strictly positive values (‘pSI’) and models that allow negative serial intervals (‘nSI’). We use the latter model throughout our presentation but highlight some effects and differences when choosing the ‘pSI’ model in Supplement 9. Due to the accumulated variability in individual disease progression and reporting, we may further assume that variance in the case interval distribution is larger than in the generation and serial interval distributions. Furthermore, the incubation period Δ_inc_ is always a positive time period, and we anticipate for the registration delay Δ_reg_ a distribution with its mass concentrated close to and right of the origin. Negative registration delays can occur if symptom onset happens after the infection was discovered. Finally, for the backward reporting offset 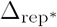 we anticipate a statistical distribution that yields mostly negative values because once a patient is discovered, the likelihood of posterior secondary infections should be minimal. The forward reporting offset 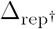 presumably is a strictly positive interval with a shape strongly influenced by the incubation period distribution. All intervals can be subject to gradual change over time. This can be attributed to the emergence of more aggressive variants of a virus, which may lead to decreased incubation periods, to the improvement of surveillance and contact tracing, which can lead to smaller reporting delays (observed in the data), or to a generally altered social contact behavior and immunity.

#### 4.1.3 Inference approach

The following equation model corresponds to the schematic in Figure 1A and brings into relation above time intervals occurring in infection pairs *AB*,

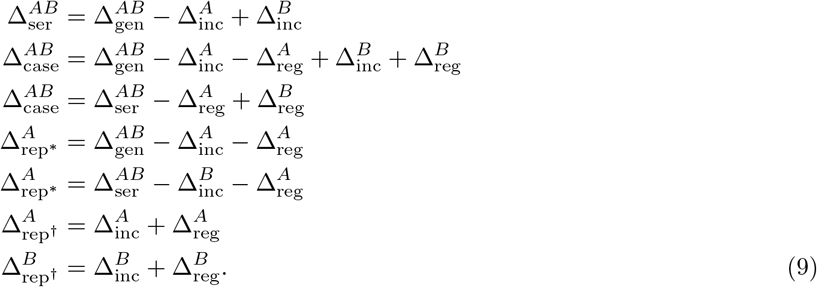

We designate the generation, serial and incubation period (Δ_gen_, Δ_ser_, Δ_inc_) as exogenous variables that can be modelled with statistical distributions found in literature (Supplement 1 and Supplement 2). We further extracted the distribution of the time between registration and symptom onset (registration delay Δ_reg_) from aggregate case data in Austria (Supplement 3 and Supplement 4). Accordingly, the remaining ‘endogenous’ variables left for inference are the case interval Δ_case_ and the forward and backward reporting offsets 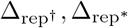. To map the equation system in a stochastic simulation framework, we differentiate between distinct instances of random variables on the left-hand side in (9) and model the difference between instances of the same variable as normal errors with a standard deviation of 3 days. We then implement the resulting expressions (Supplement 5) in a stochastic simulation framework (Python PyMC3 library [45]) and employ gradient-based MCMC algorithms to obtain stochastic samples of all random variables that comply with the formulated constraints (error terms).

We compare the obtained parameters of the fitted distributions of all exogenous variables with their prior counterparts that were found in literature and data (Supplement 6). By this we can assure that the posterior distributions only deviate marginally from the initially provided data and models while simultaneously adhering to the equation model. We further fit statistical models (skew normal distributions) to the synthetic samples of the remaining endogenous variables (case interval and reporting offset distributions) and collate their qualitative characteristics with above considerations. To honour the temporal transformation of disease intervals and weekly patterns in reporting delays, this procedure was separately performed for distinct time periods of the SARS-CoV-2 epidemic in Austria and for each day of the week. Hence, for distinct time periods and for different days of the week specific statistical models for the reporting offset distributions were obtained (compare Figure 1C).

In Supplement 7 we provide the obtained distribution parameters for all scenarios (weekday and phase of the epidemic in Austria). In Supplement 8 we visualize the obtained reporting offset distributions for all scenarios. In Supplement 9 we compare the results of our approach for real and positive serial intervals.

### 4.2 Framework for the inference of effective aggregate dispersion

#### 4.2.1 Motivation

We motivate the applied infection model with a simple formal argument, which provides an explanation as to why the variability in the number of secondary infections seems to average out in the observed numbers of daily reported cases during high-prevalence regimes.

Corresponding to the common approach for inferring reproduction dynamics (*Results*, subsection 2.4), we assume that infectious activity can be modelled with a negative binomial distribution (5),

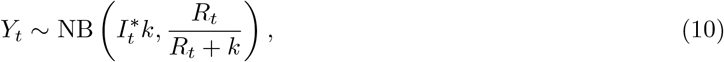

such that we obtain for the mean and variance,

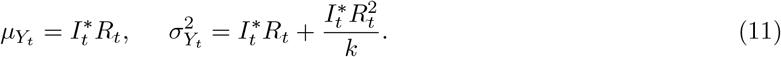

Chebyshev’s inequality yields

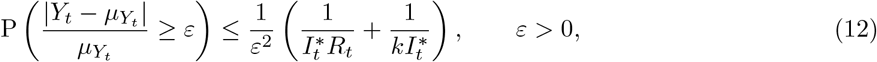

and hence, the relative deviation of infectious activity converges to zero in probability for every fixed *k*, that is

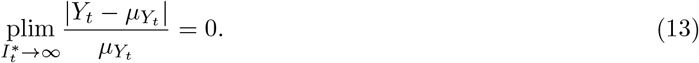

This means that, independent of the dispersion parameter *k*, the relative deviation of the daily infectious activity from the expected value converges to 0 with an increasing infectious load. This indicates that, with growing daily aggregate case numbers, the dispersion in the transmission dynamics plays a minor role and that we should in fact expect more regular time series of reported case numbers during a high-incidence regime than during a low-incidence period.

When attempting to quantify the amount of variance in a time series of aggregate reported case numbers that can be attributed to the dispersion in the number of individual secondary infections, we are faced with two major problems. First, models for the reproduction dynamics such as *Y*_*t*_ in (10) are defined by time-varying parameters that can easily change between two consecutive days. The observed time series of aggregate case numbers, however, only yields a single entry for each day. Hence, in general, we only have a single sample for a fitted model *Y*_*t*_, from which it is clearly impossible to obtain any notion of empirical variation. One way to address this issue is to assume that the parameters of a model for the reproduction dynamics of an outbreak remain unchanged for a given number of days [11]. Under this assumption, we can sample infectious load and infectious activity pairs for each day in the selected time window and measure how well a model that was fitted on these pairs explains the data observed across the respective time frame. Secondly, large parts of the variance in a time series of daily aggregate case numbers is caused by independent factors such as the individual testing behavior, the current setup of the testing regime, or the workflow of laboratories and governmental institutions. These contributions to the overall variance in the observed time series need to be separated from the variation caused by the dispersion in the number of secondary infections.

In the following, we define a model for the number of secondary infections that mitigates both of these issues and thus enables us to quantify how much dispersion at least needs to be assumed during a given period in the pandemic such that the observed variance of daily aggregate case numbers is in fact plausible.

#### 4.2.2 Statistical model for reproduction

Let *I*_*t*_ be a time series of daily aggregate case numbers, and let 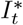 bet the corresponding time series of infectious load. We now consider a slightly modified model for the infectious activity on a day *t*, namely

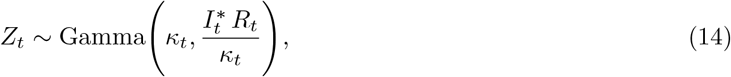

where *R*_*t*_, *κ*_*t*_ *>* 0 are the effective reproduction number and a time-dependent aggregate dispersion parameter, respectively. It holds that

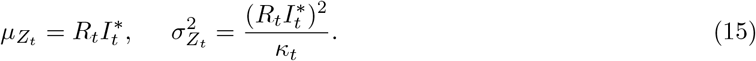

Whereas 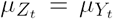, the main difference between the random variables *Z*_*t*_ and *Y*_*t*_ is how their respective standard deviations scale with the infectious load 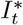. In particular, we have 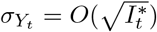 and 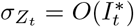 for 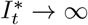. Hence, in contrast to (12), applying the Chebyshev inequality for *Z*_*t*_ to obtain

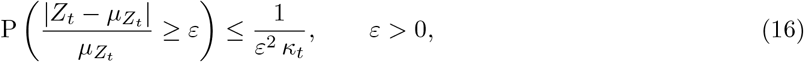

does not imply a reduction of the relative deviation from the expectation for large 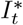. Note that this is only true when 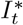 grows at least linearly with 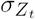, which further motivates our specific choices for the parameters in the model *Z*_*t*_.

We now consider *Z*_*t*_ as a benchmark model for the time-varying aggregate dispersion present in the incidence time series *I*_*t*_ in the following sense: If we assume that *Y*_*t*_ in (10) is an accurate model for the infection dynamics observed by *I*_*t*_ and that we can extend our model *Z*_*t*_ such that it explains most of the variance caused by independent factors such as the testing and reporting regime, it should be possible to choose the time-dependent aggregate dispersion parameter *κ*_*t*_ in the model *Z*_*t*_ increasingly large during high-incidence periods of an outbreak without making the observed data implausible. This is because, independent of the individual basic dispersion parameter *k*, the infection dynamics that yield the time series *I*_*t*_ should exhibit a relative deviation of the observed infectious activity from the expectation that goes to 0 for 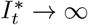 (according to (12) and (13)), while this is not necessarily true for the model *Z*_*t*_ (according to (16)). As a consequence, even with large choices for the time-varying aggregate dispersion parameter *κ*_*t*_, the model *Z*_*t*_ should be able to explain the variance observed in the time series *I*_*t*_ when 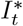 grows large.

#### 4.2.3 Inference approach

To assess the plausibility of the observed data around a day *t* given an aggregate dispersion parameter *κ*_*t*_ in the model (14), we first fit the effective reproduction numbers *R*_*t*_ on a fixed window around *t* during which we assume that the parameters describing the current infection dynamics remain constant. A typical approach in the literature is to use a constant model for *R*_*t*_ [11]. However, a constant model cannot account for variance in the data caused by daily seasonalities in the reporting or strong linear trends, both of which are usually present in daily incidence time series. Here, we will consider a model which combines a linear trend with a daily seasonal constant, namely

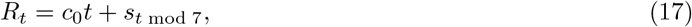

where *c*_0_, *s*_0_, …, *s*_6_ ∈ ℝ. Let here for convenience **e**_*i*_ ∈ ℝ7 for *i* ∈ {0, …, 6} denote the (*i* + 1)-th unit row vector and let 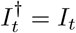 be the infectious activity on day *t* according to (3). By assuming that the parameters *c*_0_, *s*_0_, …, *s*_6_ remain constant *τ*_0_ days before and *τ*_1_ days after *t*, we define the linear problem

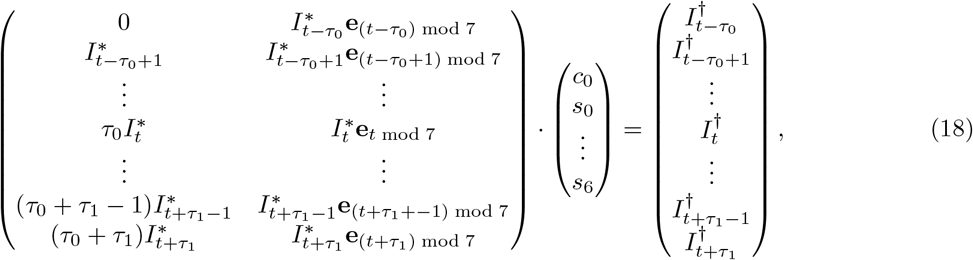

which can be solved with an ordinary least squares approach. Substituting the obtained parameters in (17) then yields a fit for the effective reproduction numbers 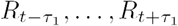 on the selected window of days around *t*. For a fixed time-dependent aggregate dispersion parameter *κ*_*t*_, which we assume to also remain constant in the period from *t − τ*_0_ to *t* + *τ*_1_, this fully defines the model *Z*_*t*_ for the selected time window. To calculate the plausibility of this model given the observed data within the selected time window, we define the following test statistic based on the squared distance from the daily expectation:

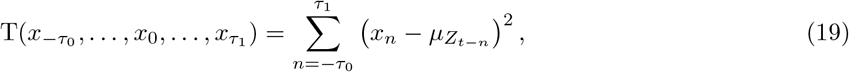

where 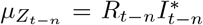. The plausibility of the model *Z*_*t*_ given the observed daily incidence time series *I*_*t*_ and a fixed parameter *κ*_*t*_ can now be assessed via the p-value that represents the probability that the test statistic (19) for samples from the random variables 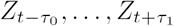 is greater or equal than the test statistic for the estimated infectious activities, that is,

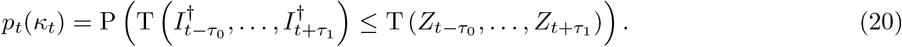

For a plausibility threshold *p* ∈ [0, 1], we write

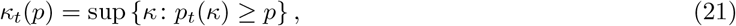

to denote the largest aggregate dispersion parameter for which the model *Z*_*t*_ is plausible given the observed daily reported cases around day *t*. The effective aggregate dispersion index (EffDI) is then defined as the square root of the reciprocal of *κ*_*t*_(*p*), that is,

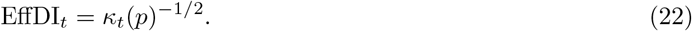

## Data Availability

Time series of reported case numbers of COVID-19 are made available by the Center for Systems Science and Engineering at Johns Hopkins University in an online repository. Parameters of the statistical distributions of disease intervals (published in literature or generated during the current study) are provided in the supplementary material with references to the sources where applicable.
Data that was used particularly in the Austrian case study was provided to us by the Austrian Agency for Health and Food Safety under a restrictive license. The results of statistical evaluations are however available in the supplementary material. Demographic data for Austria is made available by Statistics Austria.

## Data availability

Time series of reported case numbers of COVID-19 are made available by the Center for Systems Science and Engineering at Johns Hopkins University [12] in an online repository (https://github.com/CSSEGISandData/COVID-19). Parameters of the statistical distributions of disease intervals (published in literature or generated during the current study) are provided in the supplementary material with references to the sources where applicable. Data that was used particularly in the Austrian case study was provided to us by the Austrian Agency for Health and Food Safety (https://www.ages.at) [26, 27] under a restrictive license. The results of statistical evaluations are however available in the supplementary material. Demographic data for Austria is made available by Statistics Austria (https://www.statistik.at) [5].

## Code availability

The EffDI package is available on the Python Package Index (PyPI) and github (https://github.com/mdsunivie/EffDI). Additional code can be made available upon reasonable request.

## Acknowledgements

R.R. gratefully acknowledges support from the Austrian Science Fund (FWF M 2528).

## Author Contributions

G.S., L.H., R.R., N.P., and P.G. designed the study concept. L.H. and R.R. developed the EffDI and wrote the corresponding Python implementation. G.S. developed the quantifier for socio-demographic heterogeneity and the inference of the distributions of disease intervals. G.S., L.H., and R.R. conducted the numerical experiments, interpreted the results and wrote the manuscript. N.P. and P.G. supervised the study and provided input to the written text.

## Supplementary information

Supplementary material is provided in separate files.

