## Supplementary material for "Assessing the heterogeneity in the transmission of infectious diseases from time series of epidemiological data": Supplementary.pdf


#### 1 Parameters of statistical distributions of disease intervals (data)

We provide a compressed folder (`collection_interval_distributions.zip`) containing machine readable files in the JSON format with the parameters and statistics of the distributions of disease intervals found in literature. The references are listed in section 2 of the supplementary material.

#### 2 Survey on statistical distributions of disease intervals

We collected the parameters of statistical distributions of COVID-19 disease intervals reported in literature (section 1). The figures in this supplement visualize the respective probability density functions. The sources are also referenced in the main text. In the inference of the reporting offset distributions we use averaged versions of the distributions found in literature. These distributions are also contained in the compressed folder (section 1) and shown in the figures as solid lines.

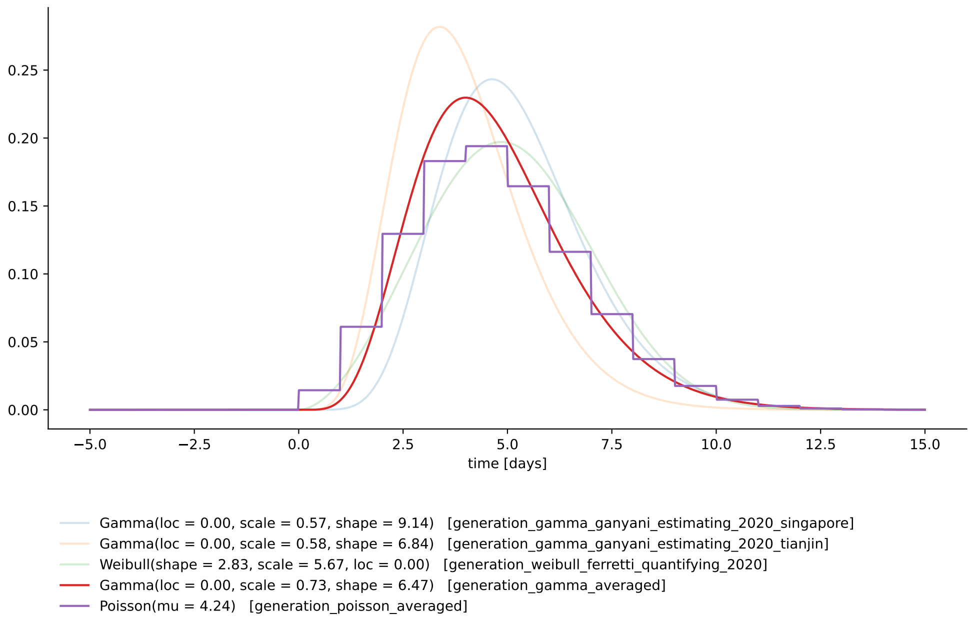


Figure 2.1: Generation interval distributions.

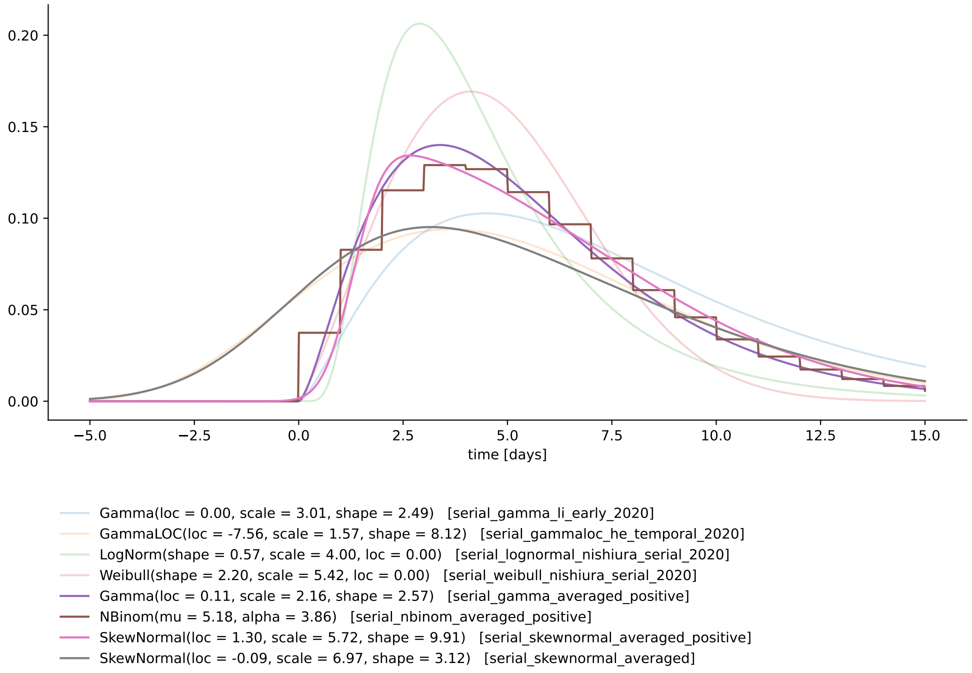


Figure 2.2: Serial interval distributions. We distinguish between models with positive support ('pSI') and models that also allow negative serial intervals ('nSI').

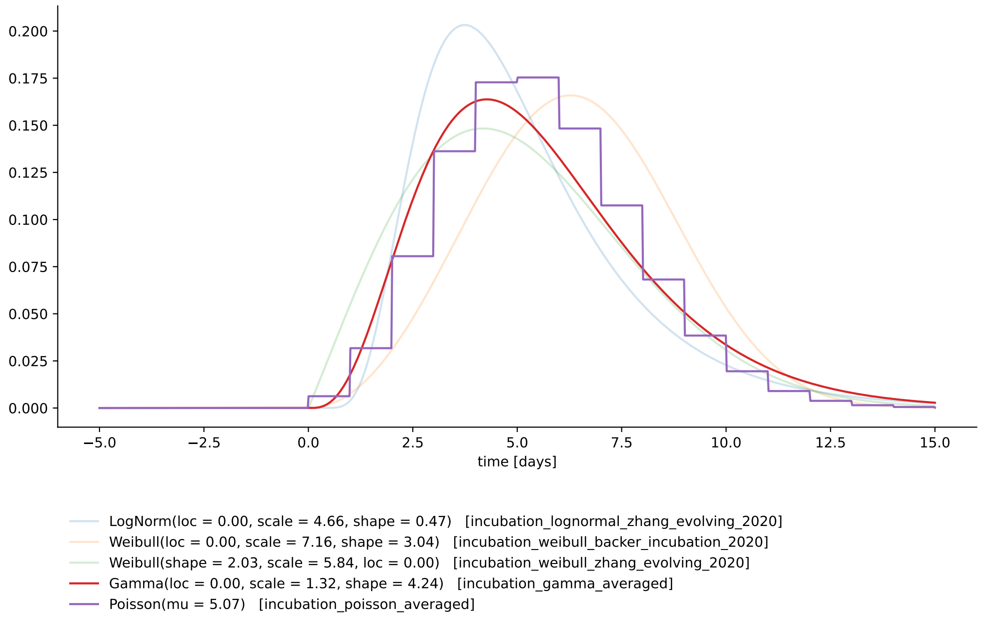


Figure 2.3: Incubation period distributions.

**nishiura_serial_2020** Nishiura, H.; Linton, N. M. & Akhmetzhanov, A. R.
Serial interval of novel coronavirus (COVID-19) infections
International Journal of Infectious Diseases, Elsevier BV, 2020, 93, 284-286
doi:10.1016/j.ijid.2020.02.060

**ferretti_quantifying_2020** Ferretti, L.; Wymant, C.; Kendall, M.; Zhao, L.; Nurtay, A.; Abeler-Dörner, L.; Parker, M.; Bonsall, D. & Fraser, C.
Quantifying SARS-CoV-2 transmission suggests epidemic control with digital contact tracing
Science, American Association for the Advancement of Science (AAAS), 2020, 368, eabb6936
doi:10.1126/science.abb6936

**ali_serial_2020** Ali, S. T.; Wang, L.; Lau, E. H. Y.; Xu, X.-K.; Du, Z.; Wu, Y.; Leung, G. M. & Cowling, B. J.
Serial interval of SARS-CoV-2 was shortened over time by nonpharmaceutical interventions
Science, American Association for the Advancement of Science (AAAS), 2020, 369, 1106-1109
doi:10.1126/science.abc9004

**du_serial_2020** Du, Z.; Xu, X.; Wu, Y.; Wang, L.; Cowling, B. J. & Meyers, L. A.
Serial Interval of COVID-19 among Publicly Reported Confirmed Cases
Emerging Infectious Diseases, Centers for Disease Control and Prevention (CDC), 2020, 26, 1341-1343
doi:10.3201/eid2606.200357

**li_early_2020** Li, Q.; Guan, X.; Wu, P.; Wang, X.; Zhou, L.; Tong, Y.; Ren, R.; Leung, K. S.; Lau, E. H.; Wong, J. Y.; Xing, X.; Xiang, N.; Wu, Y.; Li, C.; Chen, Q.; Li, D.; Liu, T.; Zhao, J.; Liu, M.; Tu, W.; Chen, C.; Jin, L.; Yang, R.; Wang, Q.; Zhou, S.; Wang, R.; Liu, H.; Luo, Y.; Liu, Y.; Shao, G.; Li, H.; Tao, Z.; Yang, Y.; Deng, Z.; Liu, B.; Ma, Z.; Zhang, Y.; Shi, G.; Lam, T. T.; Wu, J. T.; Gao, G. F.; Cowling, B. J.; Yang, B.; Leung, G. M. & Feng, Z.
Early Transmission Dynamics in Wuhan, China, of Novel Coronavirus–Infected Pneumonia
New England Journal of Medicine, Massachusetts Medical Society, 2020, 382, 1199-1207
doi:10.1056/nejmoa2001316

**zhang_evolving_2020** Zhang, J.; Litvinova, M.; Wang, W.; Wang, Y.; Deng, X.; Chen, X.; Li, M.; Zheng, W.; Yi, L.; Chen, X.; Wu, Q.; Liang, Y.; Wang, X.; Yang, J.; Sun, K.; Longini, I. M.; Halloran, M. E.; Wu, P.; Cowling, B. J.; Merler, S.; Viboud, C.; Vespignani, A.; Ajelli, M. & Yu, H.
Evolving epidemiology and transmission dynamics of coronavirus disease 2019 outside Hubei province, China: a descriptive and modelling study
The Lancet Infectious Diseases, Elsevier BV, 2020, 20, 793-802
doi:10.1016/s1473-3099(20)30230-9

**lauer_incubation_2020** Lauer, S. A.; Grantz, K. H.; Bi, Q.; Jones, F. K.; Zheng, Q.; Meredith, H. R.; Azman, A. S.; Reich, N. G. & Lessler, J.
The Incubation Period of Coronavirus Disease 2019 (COVID-19) From Publicly Reported Confirmed Cases: Estimation and Application
Annals of Internal Medicine, American College of Physicians, 2020, 172, 577-582
doi:10.7326/m20-0504

**backer_incubation_2020** Backer, J. A.; Klinkenberg, D. & Wallinga, J.
Incubation period of 2019 novel coronavirus (2019-nCoV) infections among travellers from Wuhan, China, 20–28 January 2020
Eurosurveillance, European Centre for Disease Control and Prevention (ECDC), 2020, 25
doi:10.2807/1560-7917.es.2020.25.5.2000062

**he_temporal_2020** He, X.; Lau, E. H. Y.; Wu, P.; Deng, X.; Wang, J.; Hao, X.; Lau, Y. C.; Wong, J. Y.; Guan, Y.; Tan, X.; Mo, X.; Chen, Y.; Liao, B.; Chen, W.; Hu, F.; Zhang, Q.; Zhong, M.; Wu, Y.; Zhao, L.; Zhang, F.; Cowling, B. J.; Li, F. & Leung, G. M.
Temporal dynamics in viral shedding and transmissibility of COVID-19
Nature Medicine, Springer Science and Business Media LLC, 2020, 26, 672-675
doi:10.1038/s41591-020-0869-5

**richter_schaetzung_2020** Richter, L.; Schmid, D.; Chakeri, A.; Maritschnik, S.; Pfeiffer, S. & Stadlober, E.
Schätzung des seriellen Intervalles von COVID19, Österreich
Austrian Agency for Health and Food Safety (AGES), Austrian Agency for Health and Food Safety (AGES), 2020
https://www.ages.at/en/wissen-aktuell/publikationen/schaetzung-des-seriellen-intervalles-von-covid19-oesterreich/

**ganyani_estimating_2020** Ganyani, T.; Kremer, C.; Chen, D.; Torneri, A.; Faes, C.; Wallinga, J. & Hens, N.
Estimating the generation interval for coronavirus disease (COVID-19) based on symptom onset data, March 2020
Eurosurveillance, European Centre for Disease Control and Prevention (ECDC), 2020, 25
doi:10.2807/1560-7917.es.2020.25.17.2000257

**ng_estimating_2021** Ng, S. H.-X.; Kaur, P.; Kremer, C.; Tan, W. S.; Tan, A. L.; Hens, N.; Toh, M. P.; Teow, K. L. & Kannapiran, P.
Estimating Transmission Parameters for COVID-19 Clusters by Using Symptom Onset Data, Singapore, 2020
Emerging Infectious Diseases, Centers for Disease Control and Prevention (CDC), 2021, 27, 582-585
doi:10.3201/eid2702.203018

**jhu_incubation** see lauer_incubation_2020

### 3 Parameters of reporting delay distributions (data)

We provide a compressed folder (`collection_registration_delays.zip`) containing JSON files with the parameterization of reporting delay distributions extracted from a data-set on case reporting in Austria (Austrian Agency for Health and Food Safety (AGES) – Aggregated data about symptom onset and case registration, see reference in the main text).

### 4 Visualization of delays in case reporting

Visualization of the data provided in section 3.

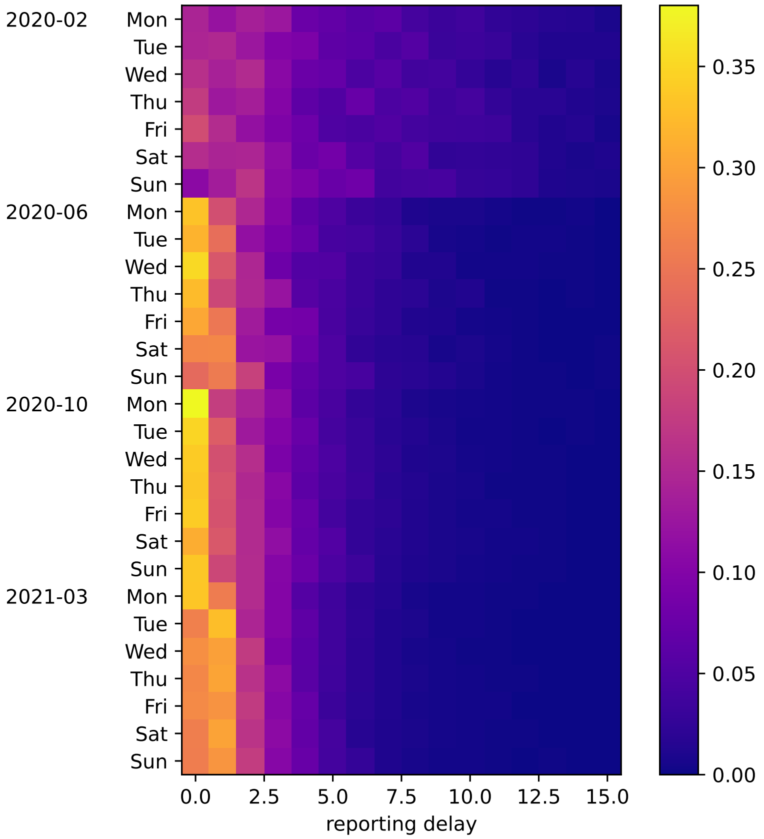


Figure 4.1: Histogram of reporting delays observed in a data-set on reported cases in Austria. Reporting delays are displayed separately for four time periods ('phases') and for each day of the week.

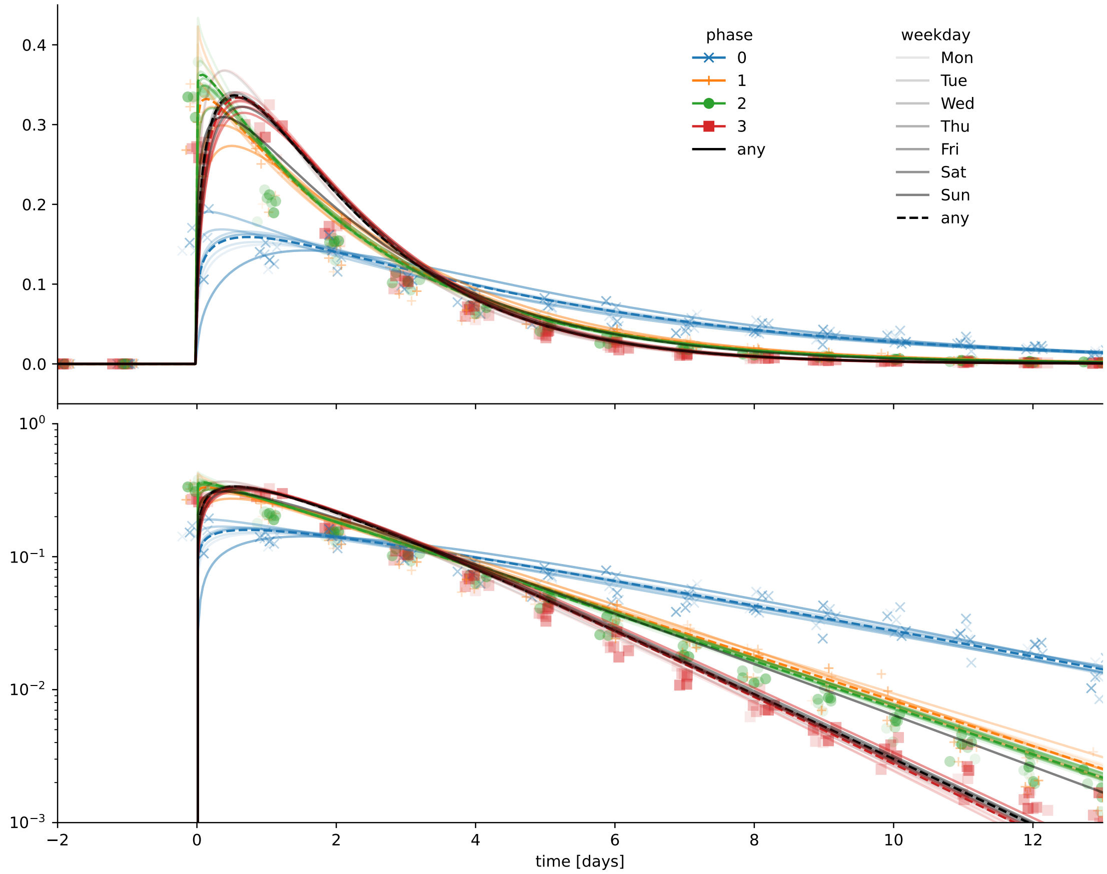


Figure 4.2: Reporting delay distributions obtained from Austrian case data. Relative frequencies (discrete distributions) in the data are shown as markers, the fitted distributions (continuous) are shown as lines. Different colors correspond to different time periods ('phases'), color intensity encodes the day of the week.

### 5 Equations and constraints for stochastic simulation

To implement the equation system developed in *Results* of the main text in a stochastic simulation framework, we differentiate between distinct instances of random variables and introduce the following intermediate definitions (deterministic combinations of 'exogenous' random variables),

$$\begin{aligned}
\Delta^{1}_{\text{ser}} &:= \Delta^{AB}_{\text{gen}} - \Delta^{A}_{\text{inc}} + \Delta^{B}_{\text{inc}}\\
\Delta^{1}_{\text{case}} &:= \Delta^{AB}_{\text{gen}} - \Delta^{A}_{\text{inc}} - \Delta^{A}_{\text{reg}} + \Delta^{B}_{\text{inc}} + \Delta^{B}_{\text{reg}}\\
\Delta^{2}_{\text{case}} &:= \Delta^{AB}_{\text{ser}} - \Delta^{A}_{\text{reg}} + \Delta^{B}_{\text{reg}}\\
\Delta^{1}_{\text{rep}^*} &:= \Delta^{AB}_{\text{gen}} - \Delta^{A}_{\text{inc}} - \Delta^{A}_{\text{reg}}\\
\Delta^{2}_{\text{rep}^*} &:= \Delta^{AB}_{\text{ser}} - \Delta^{B}_{\text{inc}} - \Delta^{A}_{\text{reg}}\\
\Delta^{A}_{\text{rep}^\dagger} &:= \Delta^{A}_{\text{inc}} + \Delta^{A}_{\text{reg}}\\
\Delta^{B}_{\text{rep}^\dagger} &:= \Delta^{B}_{\text{inc}} + \Delta^{B}_{\text{reg}}.
\end{aligned} \tag{5.1}$$

We regard the difference between instances of the same variable as stochastic errors $\varepsilon$ that are modelled as normal random variables with a standard deviation of 3 days,

$$\begin{aligned}
\varepsilon_{\text{ser}} &:= \Delta^{AB}_{\text{ser}} - \Delta^{1}_{\text{ser}}\\
\varepsilon_{\text{case}} &:= \Delta^{1}_{\text{case}} - \Delta^{2}_{\text{case}}\\
\varepsilon_{\text{rep}^*} &:= \Delta^{1}_{\text{rep}^*} - \Delta^{2}_{\text{rep}^*}.
\end{aligned} \tag{5.2}$$

### 6 Posterior distributions of the MCMC inference approach

In Figure 6.1 the parameters of the prior and posterior distributions that occurred during stochastic inference with the MCMC approach are plotted. The underlying scenario uses a serial interval distribution with strictly positive duration and a reporting delay distribution that was found for Wednesdays during the complete duration of the epidemic in Austria (filename `pSI_weekday3`). The parameters of the posterior distributions only deviate marginally from the original parameters that were retrieved from literature, indicating that the original statistical models are retained during simulation. The same can be recognized in Figure 6.2, which shows the density functions of the prior and posterior statistical models of all disease intervals. The qualitative properties of the obtained distributions of endogenous variables (reporting offset distributions and case interval) comply with the heuristic characterization of disease intervals in *Methods* in the main text.

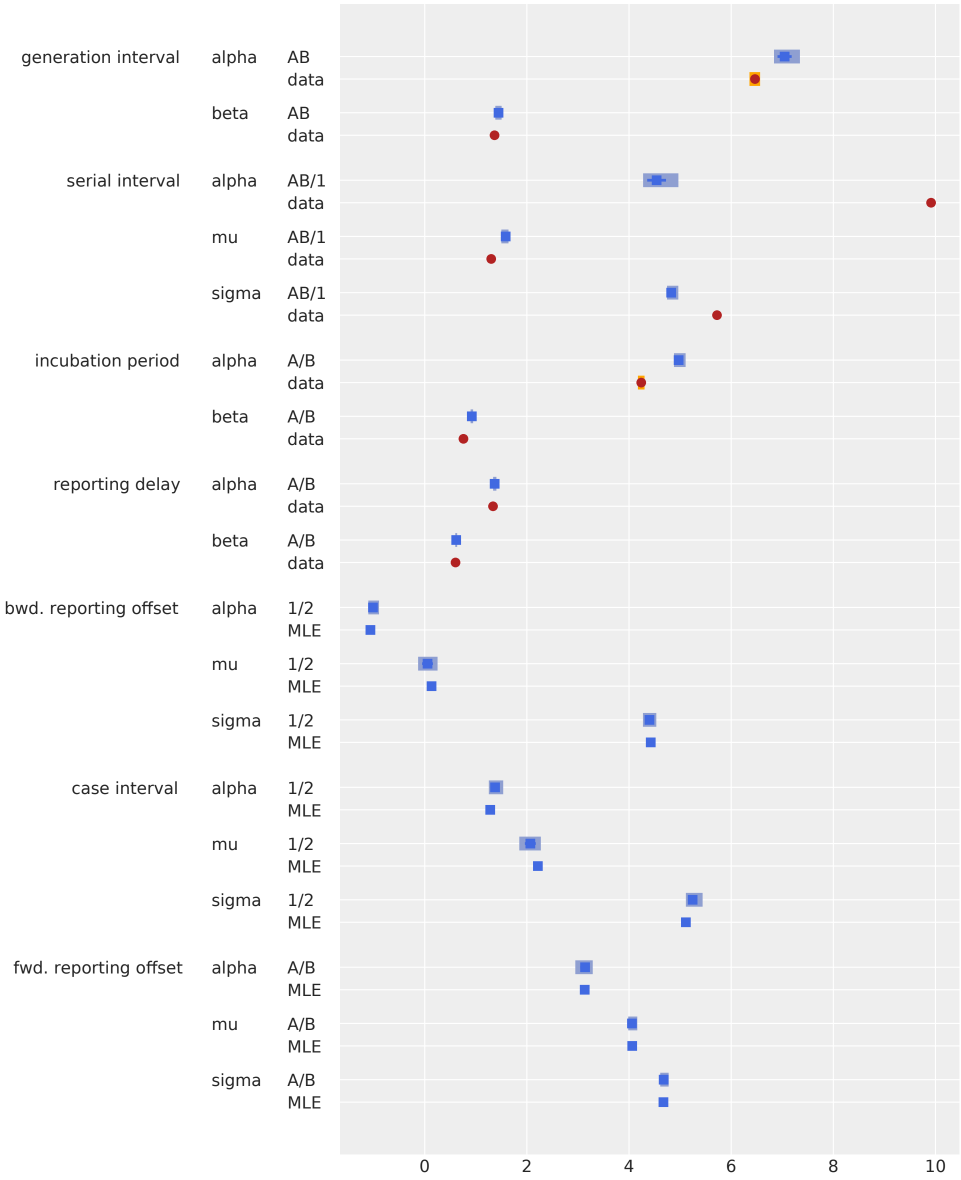


Figure 6.1: Parameters of interval distributions in the stochastic simulation approach. Prior parameters and their credible intervals (if provided by literature and data) are marked as dots in red color and labeled with 'data'. The inferred parameters and their uncertainty intervals are displayed as blue squares. In addition to the parameters obtained via MCMC sampling, we also show the maximum likelihood estimates (indicated by MLE).

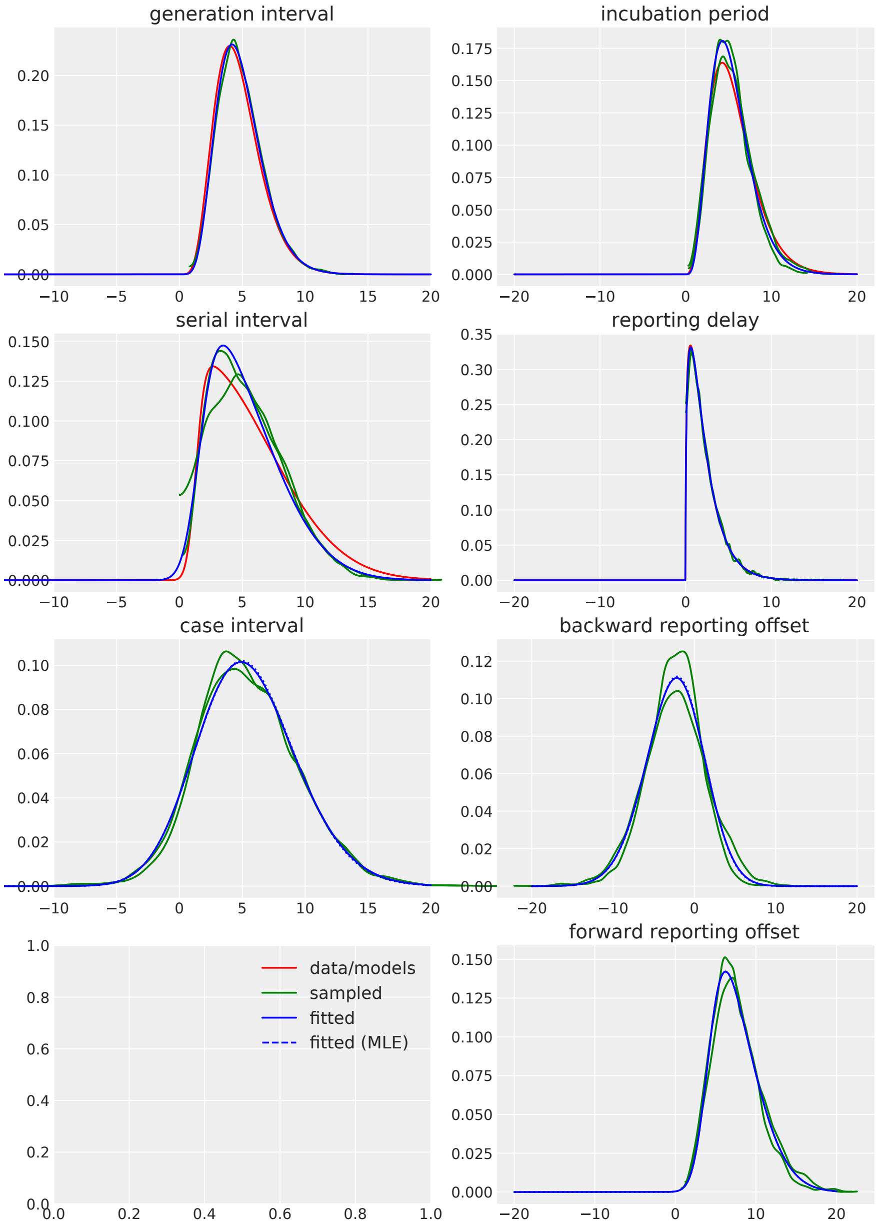


Figure 6.2: Probability density functions obtained in the stochastic simulation approach. Prior distributions that are based on literature and data (red), posterior sample distributions obtained from simulations of the constrained equation model (green); statistical models fitted to the simulation data (blue).

### 7 Parameters of the inferred distributions of disease intervals (data)

We provide a compressed folder (`collection_offset_distributions.zip`) containing JSON files with the parameters and statistics of inferred forward and backward reporting offset distributions and the inferred distributions of the case interval.

### 8 Visual display of the inferred disease interval distributions

Visualization of the data provided in section 7.

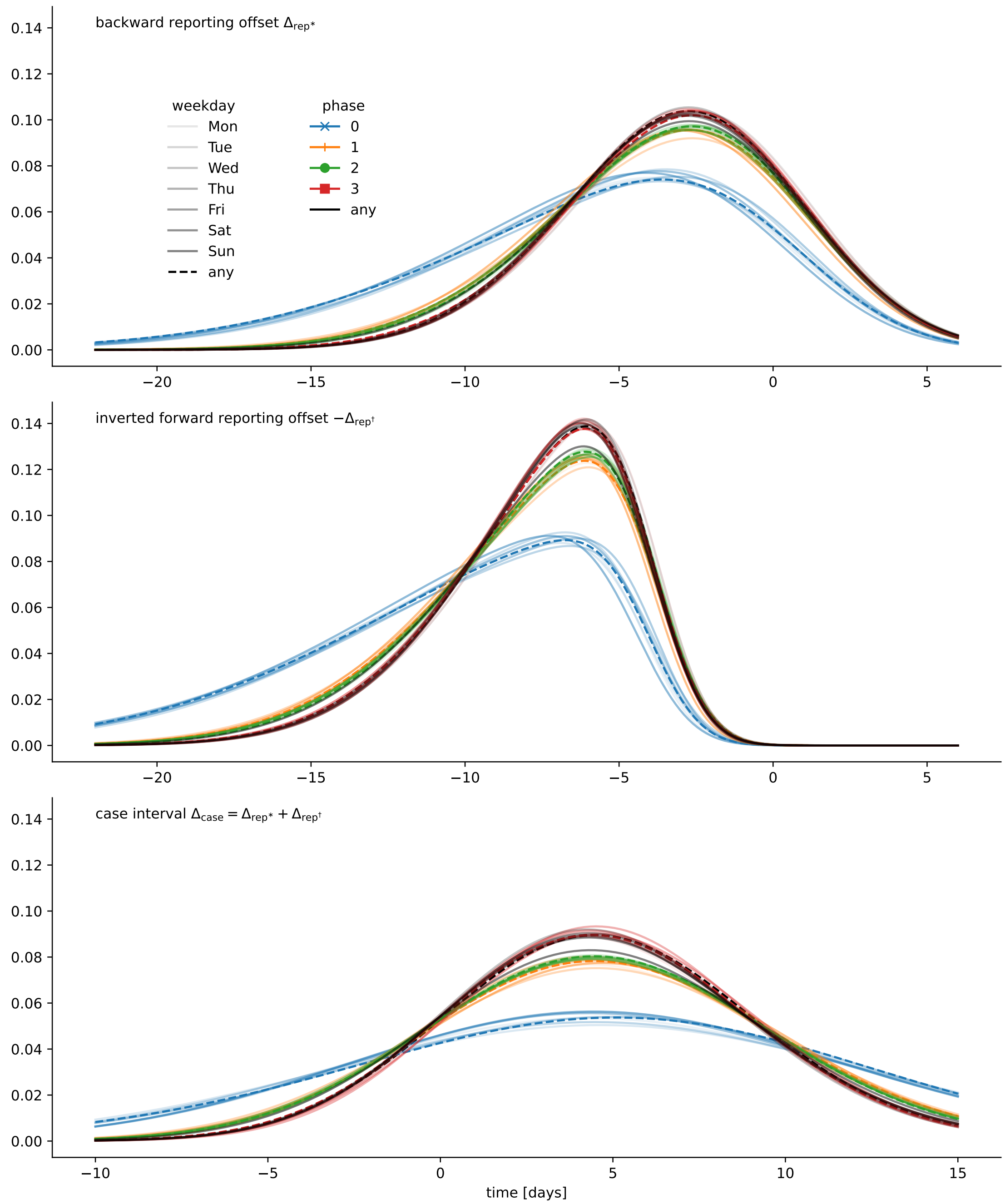


Figure 8.1: Reporting offset distributions obtained for different days of the week and for different phases of the epidemic in Austria. The displayed distributions were obtained by stochastic inference based on the serial interval model that also allows for negative duration ('nSI').

### 9 Comparison of inferred interval models under different conditions for the positivity of the serial intervals

Visual comparison of the data provided in section 7 for real and strictly positive serial intervals.

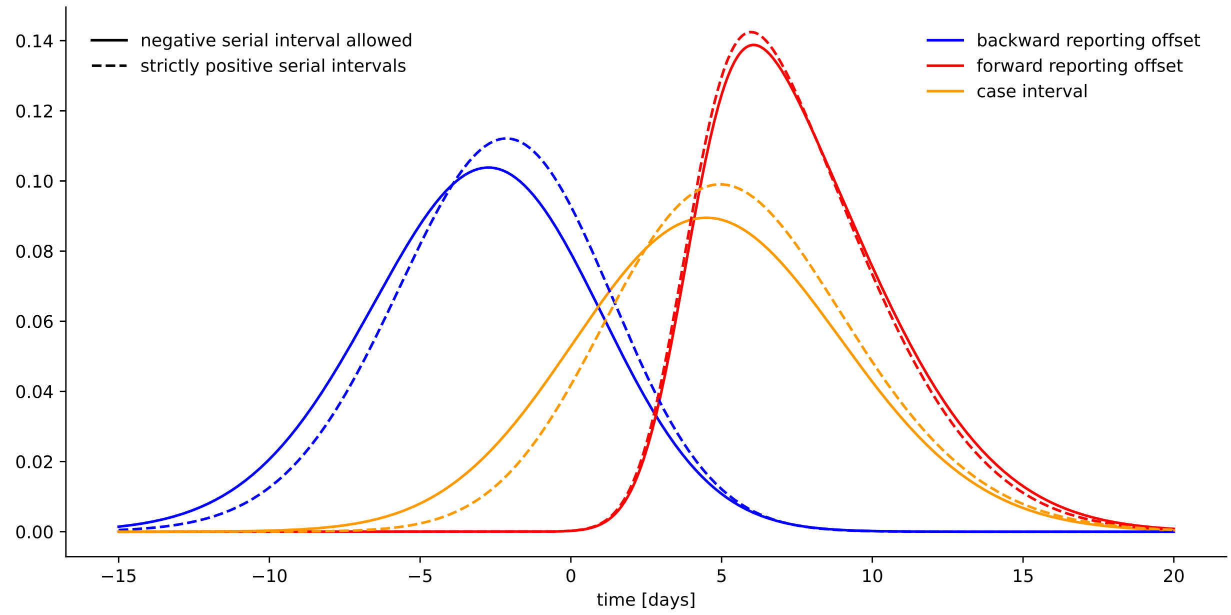


Figure 9.1: Comparison of inferred interval distributions under different conditions for the serial interval distribution ('nSI' and 'pSI').

#### 10 List of selected superspreading events in Austria

A selection of superspreading events in Austria that were reported in the news media. This list is exemplary and only serves to substantiate the quantification of socio-demographic heterogeneity presented in Figure 5 of the main text.

- C1 Geographically localized cluster in the state of Tyrol (region code 7), commonly known as the Ischgl incident. Date: 2020-03 References: https://www.politico.eu/article/the-austrian-ski-town-that-spread-coronavirus-across-the-continent/, https://web.archive.org/web/20200323121016/https://www.politico.eu/article/the-austrian-ski-town-that-spread-coronavirus-across-the-continent/
- C2 Reports about infection clusters in retirement homes and resulting from social conventions in Vienna and Upper Austria. Date: 2020-03-22 – 2020-03-25 References: https://ooe.orf.at/stories/3040384/, https://kurier.at/chronik/wien/coronavirus-ausbruch-in-vier-wiener-pensionistenheimen/400792283
- C3 Multiple larger SSEs (60 infected) in logistic centers and in social amenities in and around Vienna (region code 9). Date: 2020-05-04 – 2020-05-18 References: https://wien.orf.at/stories/3049245/
- C4 Multiple larger SSEs in the state of Lower Austria (region code 3). 26 cases in religious event and 40+ cases ins meat factory. Date: 2020-07-17 References: https://noe.orf.at/stories/3058053/
- C5 Localized cluster of young and middle-aged persons in tourist destination in St. Wolfgang, Upper Austria (region code 4). 62 infected directly attributed to SSE, 110 secondary infections estimated. Date: 2020-07-27 References: https://www.derstandard.de/story/2000119006698/st-wolfgang-nimmt-den-cluster-gelassen
- C6 Reports about infection clusters during private meetings and celebration. Date: 2020-08-14 – 2020-08-18 References: https://www.diepresse.com/5852823/282-neuinfektionen-in-osterreich-die-alterspyramide-hat-sich-vollig-verandert, https://www.sn.at/salzburg/chronik/der-naechste-cluster-in-salzburg-16-covidfaelle-in-einrichtung-der-lebenshilfe-91666543
- C7 SSEs (20 to 50 detected cases) in public events and entertainment across the country. Date: 2020-09-14 – 2020-09-15 References: https://www.vienna.at/coronavirus-cluster-an-muk-privatuni-in-wien-auf-46-infizierte-erhoeht/6740633, https://tirol.orf.at/stories/3067024/
- C8 Reports about infection clusters (50+ infected) in retirement homes. Date: 2021-01-11 – 2021-01-14 References: https://kurier.at/chronik/wien/virus-mutation-kam-wahrscheinlich-mit-einem-mitarbeiter/401155377 https://salzburg.orf.at/stories/3084397/

### 11 Additional details on the statistical models for reproduction

This is to provide some additional details on the statistical models for reproduction in (4), (5) and (6) in the main text. We use the following parameterizations of probability distributions,

$$\text{NB}(\text{number of failure until stop}, \text{probability of success})$$
$$\text{Poisson}(\text{rate})$$
$$\text{Gamma}(\text{shape}, \text{scale}).$$

The mean and the variance of the negative binomial distribution and the gamma distribution agree for the parameter choices

$$\text{NB}(r, p) \qquad \text{and} \qquad \text{Gamma}\left(rp, \frac{1}{1-p}\right).$$

Let $X_t$ bet the expected number of secondary cases produced by the infectious population $I_t^*$. A common approach for a statistical model for infectious activity is

$$I_t^\dagger \sim \text{Poisson}(X_t). \tag{11.1}$$

$X_t$ is often modelled as the sum of individual gamma distributed reproduction factors $R_i$ such that

$$I_t^\dagger \sim \text{Poisson}\left(\sum_{i=1}^{I_t^*} R_i\right), \qquad R_i \sim \text{Gamma}(k, R_t/k), \tag{11.2}$$

which, as a gamma-Poisson mixture, can be written as (equation (4) in the main text),

$$I_t^\dagger \sim \text{NB}\left(k\, I_t^*, \frac{R_t}{R_t + k}\right), \tag{11.3}$$

where by the summation property of the gamma distribution

$$X_t = \sum_{i=1}^{I_t^*} R_i \sim \text{Gamma}(I_t^* k, R_t/k). \tag{11.4}$$

Rewriting

$$X_t = I_t^* \left(\frac{1}{I_t^*} \sum_{i=1}^{I_t^*} R_i\right) \tag{11.5}$$

and replacing the average individual reproduction factor with $\bar{R}_t \sim \text{Gamma}(k, R_t/k)$, we obtain by the scaling property of the gamma distribution,

$$X_t = I_t^* \bar{R}_t \sim \text{Gamma}(k, I_t^* R_t/k). \tag{11.6}$$

The resulting gamma-Poisson mixture for infectious activity is

$$I_t^\dagger \sim \text{NB}\left(k, \frac{I_t^* R_t}{I_t^* R_t + k}\right). \tag{11.7}$$

For a broad range of configurations this model can be approximated by

$$I_t^\dagger \sim \text{NB}\left(\frac{I_t^* R_t k}{I_t^* R_t - k}, \frac{I_t^* R_t - k}{I_t^* R_t}\right), \tag{11.8}$$

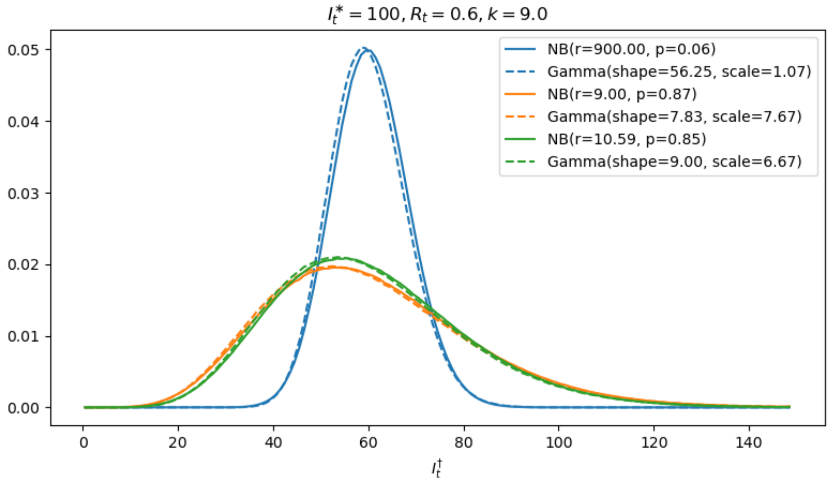


Figure 11.1: Densities of the models in Table 11.1 for a choice of parameters. The order in the legend of the figure corresponds to the order in the table.

which, in turn, can be approximated by (equation (6) in the main text)

$$I_t^\dagger \sim \text{Gamma}\left(k, \frac{I_t^* R_t}{k}\right). \tag{11.9}$$

The resulting gamma model has a comparably simple parameterization and statistical properties and satisfies the considerations in the paper. In Table 11.1 we compare the parameters and main statistical properties of the models discussed above.

| model | mean | variance | coefficient of dispersion | coefficient of variation |
|---|---|---|---|---|
| $I_t^\dagger \sim \text{NB}\left(k\, I_t^*, \frac{R_t}{R_t+k}\right)$<br>$I_t^\dagger \sim \text{Gamma}\left(\frac{k I_t^* R_t}{R_t+k}, \frac{R_t+k}{k}\right)$ | $I_t^* R_t$ | $k^{-1} I_t^* R_t^2 + I_t^* R_t$ | $k^{-1} R_t + 1$ | $k^{-1/2}\sqrt{\frac{R_t+k}{I_t^* R_t}}$ |
| $I_t^\dagger \sim \text{NB}\left(k, \frac{I_t^* R_t}{I_t^* R_t+k}\right)$<br>$I_t^\dagger \sim \text{Gamma}\left(\frac{k I_t^* R_t}{I_t^* R_t+k}, \frac{I_t^* R_t+k}{k}\right)$ | $I_t^* R_t$ | $k^{-1}(I_t^* R_t)^2 + I_t^* R_t$ | $k^{-1} I_t^* R_t + 1$ | $k^{-1/2}\sqrt{\frac{I_t^* R_t+k}{I_t^* R_t}}$ |
| $I_t^\dagger \sim \text{NB}\left(\frac{I_t^* R_t k}{I_t^* R_t-k}, \frac{I_t^* R_t-k}{I_t^* R_t}\right)$<br>$I_t^\dagger \sim \text{Gamma}\left(k, \frac{I_t^* R_t}{k}\right)$ | $I_t^* R_t$ | $k^{-1}(I_t^* R_t)^2$ | $k^{-1} I_t^* R_t$ | $k^{-1/2}$ |

Table 11.1: Comparison of statistical models for reproduction.

### 12 Parameter variation studies

#### 12.1 Variation of the mode in the pre-processing of reported case numbers

This supplement investigates the impact of different disease interval models on EffDI. We continuously transform the 'regularizing' model (equation (2) in the main text) into the 'raw' model (equation (3) in the main text), which is commonly used for inferring effective reproduction numbers. This is achieved by transforming the parameters (shape, location, scale) of the corresponding disease interval distributions according to

$$\Delta_{\text{rep}^\dagger} \longleftrightarrow \delta_0 \tag{12.1}$$

$$\Delta_{\text{rep}^*} \longleftrightarrow \Delta_{\text{rep}^*} + \Delta_{\text{rep}^\dagger} = \Delta_{\text{case}} \approx \Delta_{\text{ser}} \approx \Delta_{\text{gen}}. \tag{12.2}$$

The transition of the (continuous versions of the) interval distributions is visualized in Figure 1 in the main text and here in Figure 12.1. The parameters $\gamma$ and $\delta$ are used as scaling parameters. On the right-hand side, the delta distribution $\delta_0$ yields the raw reported case numbers as the time series of infectious activity, whereas infectious load is obtained as the convolution with the case interval distribution. Because $\mathbb{E}[\Delta_{\text{rep}^*}] + \mathbb{E}[\Delta_{\text{rep}^\dagger}] = \mathbb{E}[\Delta_{\text{case}}]$, the characteristic inter-case period is retained during the transition.

We see in Figure 12.1 that the resulting effective reproduction number is affected only marginally by this transition. However, the obtained EffDI scales towards a homogeneous progression (less distinctive changes in amplitude) with lower detected stochasticity when the 'regularizing' model is approached. This confirms that the regularization of the time series of load and activity impedes the assessment of stochasticity and effective aggregate dispersion in reproduction dynamics.

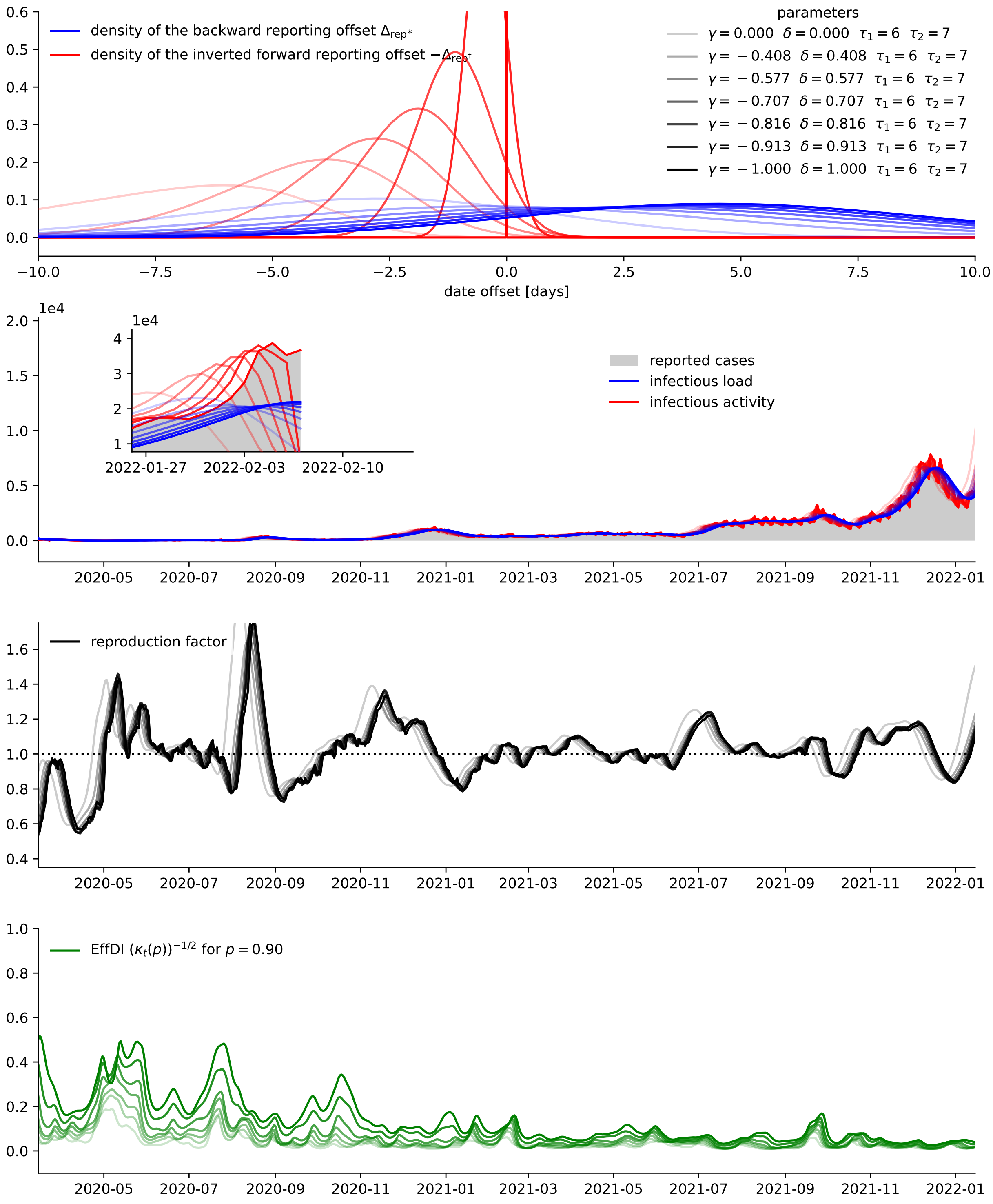


Figure 12.1: Transition between the 'raw' model for load and activity and the 'regularizing' model (equations (2) and (3) in *Results*). Publicly available data on reported case numbers provided by the Johns Hopkins University Center for Systems Science and Engineering (see reference in the main text) for the SARS-CoV-2 pandemic in South Korea was used. To avoid truncation errors the results were clipped.

#### 12.2 Variation of the shape of disease interval distributions

We show that the calculation of a reproduction factor and of EffDI is not sensitive to the shape of the used disease interval distributions. In Figure 12.2 the shape parameter of the backward reporting offset distribution was modulated while holding the mean and variance fixed. For different shapes of the reporting offset, the resulting effective reproduction factor and the EffDI only change marginally. Leaving aside the technical background of the convolution operation, this indicates that for our application merely the mean and variance of disease intervals (backward and forward reporting offset, case interval) are relevant for determining infectious load and activity.

We recapitulate that the distance between the expectation values $\mathbb{E}[\Delta_{\text{rep}^*}] - \mathbb{E}[-\Delta_{\text{rep}^\dagger}] = \mathbb{E}[\Delta_{\text{case}}] \approx \mathbb{E}[\Delta_{\text{ser}}] \approx \mathbb{E}[\Delta_{\text{gen}}]$ controls the inter-case period and that the variance (and abruptness) of the convolution kernels controls the regularization of load and activity. Simultaneous translation of both expectation values, however, results in a shift of the obtained effective reproduction factor and EffDI. In the perspective of real-time assessment and for other applications it might be necessary to exactly model certain properties such as the strictly positive range of disease intervals.

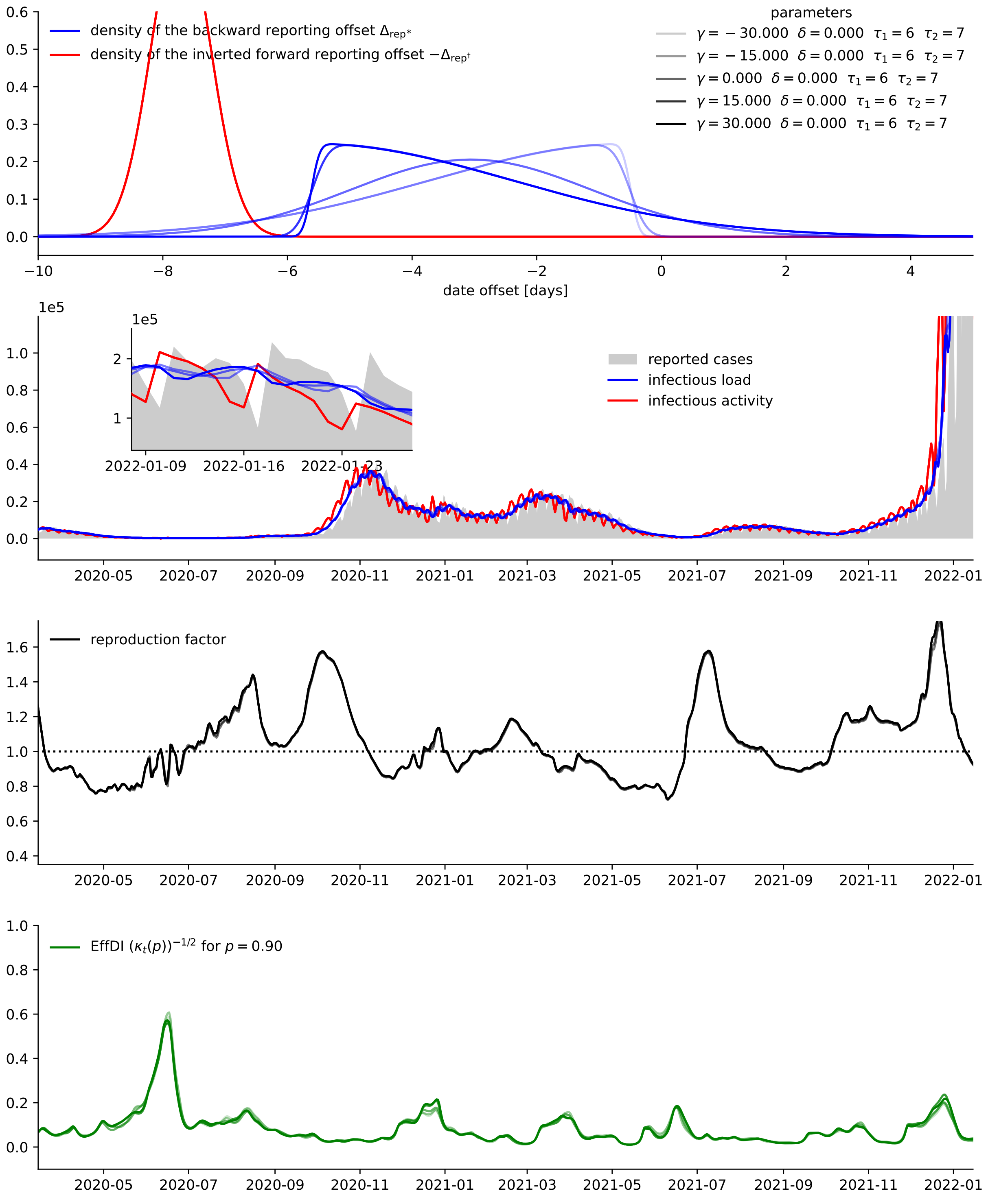


Figure 12.2: Variation of the shape of the backward reporting offset distribution. Publicly available data on reported case numbers provided by the Johns Hopkins University Center for Systems Science and Engineering (see reference in the main text) for the SARS-CoV-2 pandemic in Italy was used. To avoid truncation errors the results were clipped.
